# Green space and loneliness: a systematic review with theoretical and methodological guidance for future research

**DOI:** 10.1101/2022.05.13.22275038

**Authors:** Thomas Astell-Burt, Terry Hartig, I Gusti Ngurah Edi Putra, Ramya Walsan, Tashi Dendup, Xiaoqi Feng

## Abstract

Urban greening may help to reduce the population health impacts of loneliness and its concomitants, such as hopelessness and despair. However, the literature lacks both a critical appraisal of extant evidence and a conceptual model to explain how green space would work as a structural intervention. Both are needed to guide decision making and further research. We conducted a systematic review of quantitative studies testing associations between green space and loneliness, searching seven databases. Twenty two studies were identified by 25/01/2022. Most of the studies were conducted in high-income countries and fifteen (68%) had cross-sectional designs. Green space was measured inconsistently using either objective or subjective indicators. Few studies examined specific green space types or qualities. The majority of studies measured general loneliness (e.g. using the UCLA loneliness scale). Different types of loneliness (social, emotional, existential) were not analysed. Of 132 associations, 88 (66.6%) indicated potential protection from green space against loneliness, with 44 (33.3%) reaching statistical significance (p<0.05). We integrated these findings with evidence from qualitative studies to elaborate and extend the existing pathway domain model linking green space and health. These elaborations and extensions acknowledge the following: (a) different types of green space have implications for different types of loneliness; (b) multilevel circumstances influence the likelihood a person will benefit or suffer harm from green space; (c) personal, relational, and collective processes operate within different domains of pathways linking green space with loneliness and its concomitants; (d) loneliness and its concomitants are explicitly positioned as mediators within the broader causal system that links green space with health and wellbeing. This review and model provide guidance for decision making and further epidemiological research on green space and loneliness.

## INTRODUCTION

Many scientists and health practitioners warn of an epidemic of loneliness affecting up to a quarter of adults in countries such as the US,^1^ the UK,^2^ Australia^3^ and Sweden.^4^ Loneliness, characterised by felt deprivation of connection, comradery and companionship, is a concept often misunderstood and misconstrued. Loneliness is stereotypically associated with ageing, yet it can affect people of any age.^5^ It is a highly sensitive, often stigmatised condition^6^ described alarmingly by some commentators as ’a social cancer’^7^ and ’the leprosy of the 21st century’.^8^ Loneliness is typically overlooked by health sector-led prevention strategies and yet, scientists and health practitioners now understand it to be an aversive state associated with an increased risk of multiple chronic diseases.^9–15^ Loneliness is not a disease, but it has been medicalised.^16^ Attempts to address loneliness so far have been mostly person-focussed and weak, or ineffective.^17 18^

Policy options that shift the locus of intervention from individuals to the community context need to be identified.^19^ Urban greening was specifically highlighted as a policy option in the UK loneliness strategy.^20^ The potential of parks and other forms of green space to be part of a scalable public policy strategy to reduce loneliness is highly compelling, especially in light of the already well-documented benefits for health,^21–23^ climate and biodiversity.^24–26^ Recognizing these other benefits, cities around the world have made durable commitments to increase vegetation cover and quality (e.g. Sydney,^27^ Canberra,^28^ Barcelona,^29^ Seattle,^30^ Singapore^31^ and Vancouver^32^). These may be amenable to tailoring with a view to ameliorating loneliness.

It is important to recognize that ‘loneliness’ is often used in a general sense, but previous work (e.g. Weiss^33^) has distinguished between ‘social loneliness’ and ‘emotional loneliness’. Whereas the former refers to the feeling of being marginalized from a network of friends and family, the latter occurs when a person feels deprived of significant others whom they feel they could rely on, or share intimate moments with. A third way of feeling lonely, ‘existential loneliness’, involves a sense of emptiness arising from feelings of disconnection and disempowerment.^34^ Despair is a close companion of existential loneliness, and loneliness in general. It is described as having multiple dimensions including ‘cognitive’ (feelings of defeat, worthlessness and hopelessness), ‘emotional’ (excessive sadness, hostility and anhedonia), ‘behavioural’ (risk taking, recklessness, self-destructiveness), and ‘biological’ (homeostatic imbalance).^35^ Case and Deaton attributed rising ‘deaths of despair’ to multiple processes aligned with loneliness that have “cumulatively undermined the meaning of life”.^36^ It is plausible that different types and qualities of green spaces afford different experiences and so may work to reduce different forms of loneliness and its concomitants.

Numerous qualitative studies^37–41^ and theoretical contributions^42 43^ indicate multiple potential pathways by which green space may reduce loneliness, both in general and in people with particular life circumstances. However, there is currently no model coherently weaving together these rich seams of scholarship. Likewise, the literature lacks a review of quantitative studies that estimate association between green space and loneliness, whether approached as direct effects or as indirect effects realized through mediating processes. Accordingly, this paper reports findings from a systematic review of the quantitative research that provides estimates of association and mediating processes. We integrate these quantitative findings with findings from a selective review of qualitative studies in a conceptual model that provides needed theoretical and methodological guidance for future investigation. The model and the results it organizes will be useful to social policy makers, urban planners and landscape architects who can use placemaking and greening strategies to help reduce levels of loneliness in society, while also pursuing other sustainability goals, including climate change adaptation and biodiversity protection.

## METHODS

### Search strategy

This systematic review followed the guidelines from the Preferred Reporting Items for Systematic Review and Meta-Analysis (PRISMA) ^44^. The systematic search was conducted on 25 January 2022 using seven frequently accessed databases. These include PubMed, Scopus, Web of Science, PsycINFO, CINAHL, Cochrane Library, and ProQuest. Previously published systematic reviews guided identification and selection of search terms relevant to green space^45^ and loneliness.^10^ Table 1 presents the terms that were searched in the titles, abstracts, and/or keywords of the articles. Moreover, the systematic search also included checking the references from eligible articles.

**Table 1.** Search terms used for the systematic search

| Main keywords | Search terms |
| --- | --- |
| Green space | “green space” OR greenspace OR “green area” OR greenness OR greenery OR grass OR tree OR natur* OR “natural environment” OR vegetation OR park* OR “open space” OR garden OR “national park” OR “play space” OR “urban park” OR “recreation* resource” OR woodland OR wilderness OR “wild land” OR “natural land” OR “municipal land” OR “community land” OR “public land” OR “open land” OR “shinrin-yoku” OR “forest bathing” OR “park availability” OR “city park” OR bush |
| Loneliness | lonely OR loneliness OR lone* OR isolation OR “social isolation” OR “feeling isolated” OR “solitude” OR “solace” OR “seclusion” OR “lives alone” OR “living alone” OR “felt alone” OR “feeling alone” OR “social withdrawal” OR “socially disconnected” |

*\*truncation symbol used to search all possible variations of the word*

### Eligibility or selection criteria

The selection criteria specified studies that: 1) used quantitative methods with an observational or experimental design; 2) assessed at least one measure of green space in relation to loneliness; 3) utilised either objective or subjective/perceived measures of green space; and 4) examined loneliness as an outcome or a mediating variable through which green space affected some other health outcome. Further, the studies selected for review were published 5) since 2000; 6) in peer-reviewed journals; 7) in English. Non-peer reviewed articles, commentaries, case reports and conference papers, studies that did not test associations, and studies examining proxy measures of loneliness, such as living alone and marital status, were excluded from the systematic review, but were retained to help inform discussion of future areas for quantitative research.

The main outcome of interest in this review was loneliness. Given the association, though not direct equivalence, of loneliness with social isolation, terms such as social isolation, social withdrawal, and social disconnectedness were also included to ensure a comprehensive search of the literature. The main independent variable was green space. Green space refers to both natural and artificial (designed and built) outdoor green and open spaces with prominent vegetation components such as trees (including street trees), shrubs, grass and flowerbeds. It includes gardens, parks and diverse other settings that people can view or visit.^23^ Green space in this review includes all attributes and features outlined in Table 1. Green space indicators assessed using land use databases, geographic information systems (GIS), satellite imagery, and field observations were regarded as objective measures. Exposure variables obtained through interviews and questionnaires were classified as subjective measures.

### Selection strategy and data extraction

The process to search and select articles for this systematic review is illustrated in Figure 1. All articles retrieved from each of the databases were downloaded into the reference manager EndNote. Duplicate papers were removed initially by using the EndNote function followed by manual removal. The titles and abstracts were assessed by two reviewers independently against the selection criteria (EP, TD). Each reviewer then reviewed the articles requiring full-text assessment. Any disagreements and differences were resolved through discussion and consultation with a third reviewer (RW). Data on the publication year, author, study design, study sample and size, exposure measure and assessment, outcome measure, the measure of association, and covariates adjusted for were extracted (Supplementary Table 1).

**Figure 1:**
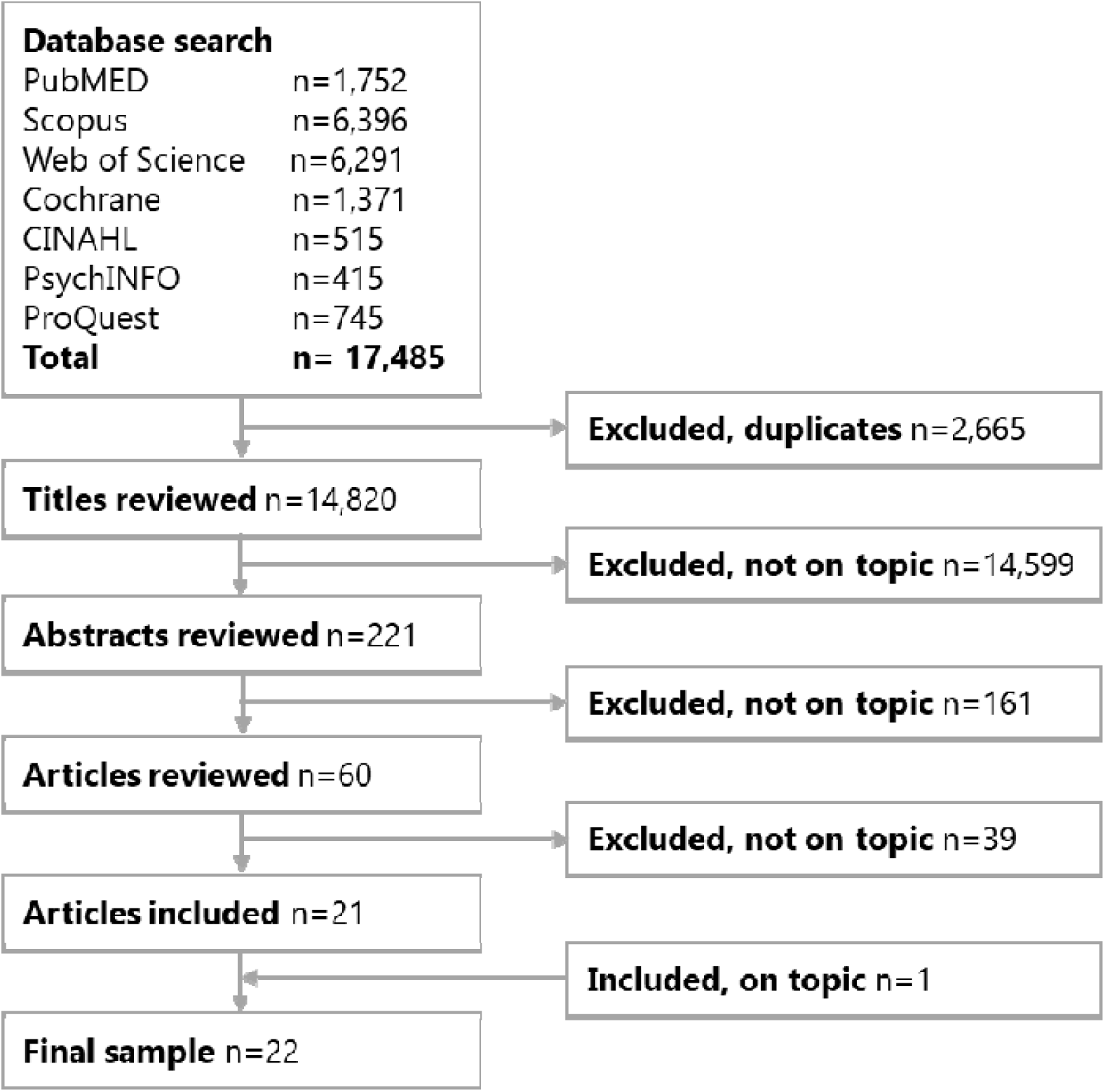
Flowchart illustrating the systematic search process

### Data analysis

The risk of bias and quality of each study included were assessed by two reviewers (EP, TD) using the U.S. National Institutes of Health quality assessment tool for intervention and observational (cohort and cross-sectional) studies.^46^ Two reviewers assessed the quality of eligible papers and any discrepancies were discussed with the third reviewer (RW). The evidence, including direction and magnitude of association in the selected studies, was narratively synthesised. Meta-analysis was not possible due to heterogeneity in study designs and variable measurement. The findings were then discussed and potential areas of future research were proposed.

## RESULTS

### Sample

Figure 1 presents the results of systematic search using the PRISMA guidelines. Out of the total of 17,485 articles retrieved from the seven databases, 2,665 duplicates were removed, followed by the exclusion of 14,559 articles that did not have information on green space and/or loneliness, leaving 221 articles for abstract review. After abstract and full-paper review, a total of 22 papers were included.

### Study characteristics

Table 2 and Supplementary Table 1 present a summary of the studies included in the systematic review. The majority (13 studies) were conducted in European countries: five in the Netherlands^47–51^; three from the UK^52–54^; one each in Spain^55^ and Germany^56^; and three that used data from multiple European countries.^57–59^ The remaining studies were conducted in the US (four studies),^60–63^ Australia (1),^64^ China (1),^65^ Hong Kong (1),^66^ Japan (1),^67^ and other multiple countries (1).^68^ Of the 22 studies, three were randomized trials,^55 60 63^ two were small-scale quasi-experiments with longitudinal (pre/post) designs,^62 66^ two were longitudinal studies,^64 68^ and the remaining 15 were cross-sectional surveys. The unit of analysis in all of these studies was the individual, with no ecological studies examining rates of loneliness across geographical units observed. Two^64 68^ and thirteen^48 49 53–60 63 66 67^ of the studies were judged to be of good and fair quality, respectively while the remaining seven studies^47 50–52 61 62 65^ had poor quality. Around 82% of the studies were conducted in the most recent 5-year period (2016-2021). Data collection for five studies was carried out during the Covid-19 pandemic.^53 59 63 65 67^

**Table 2.**
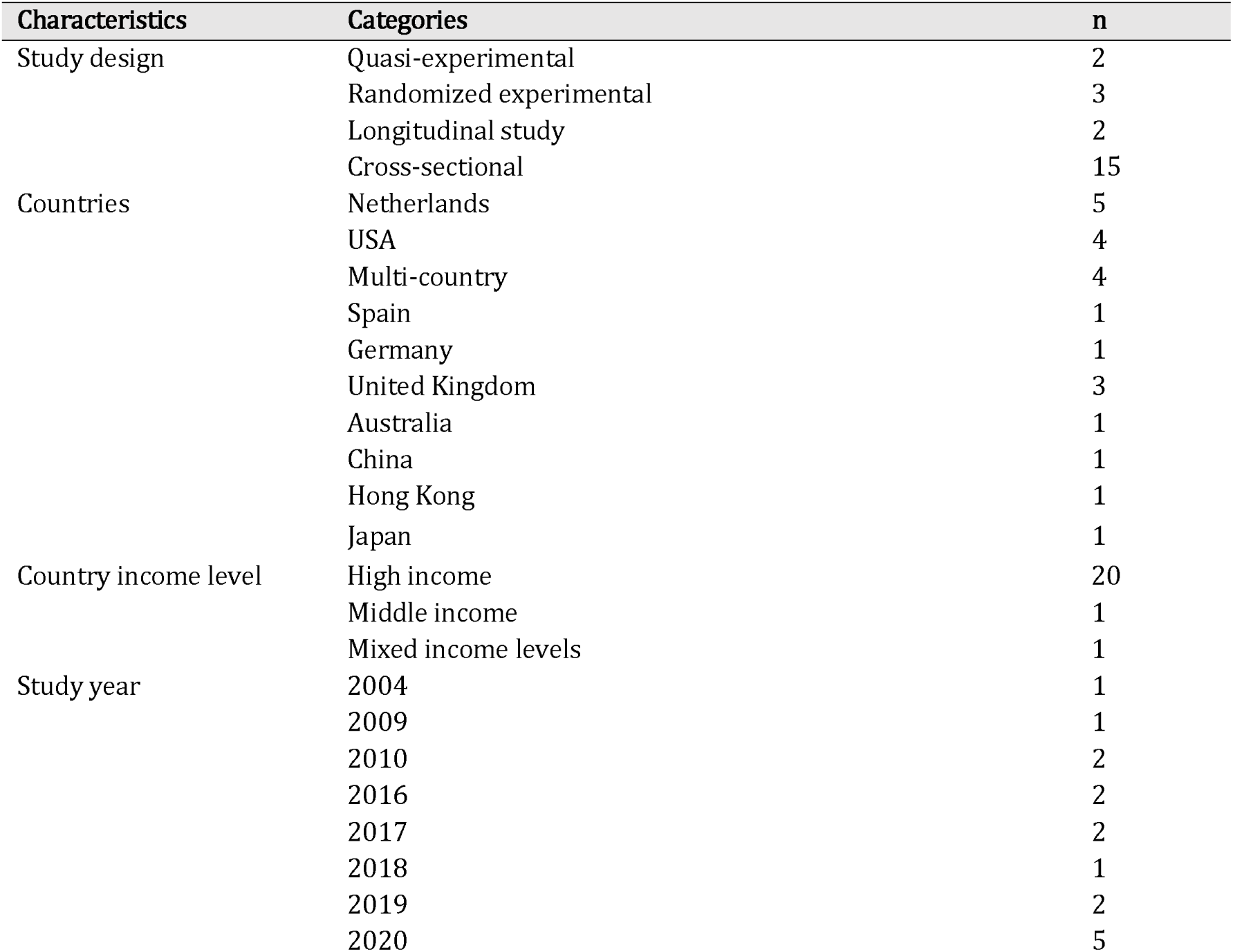
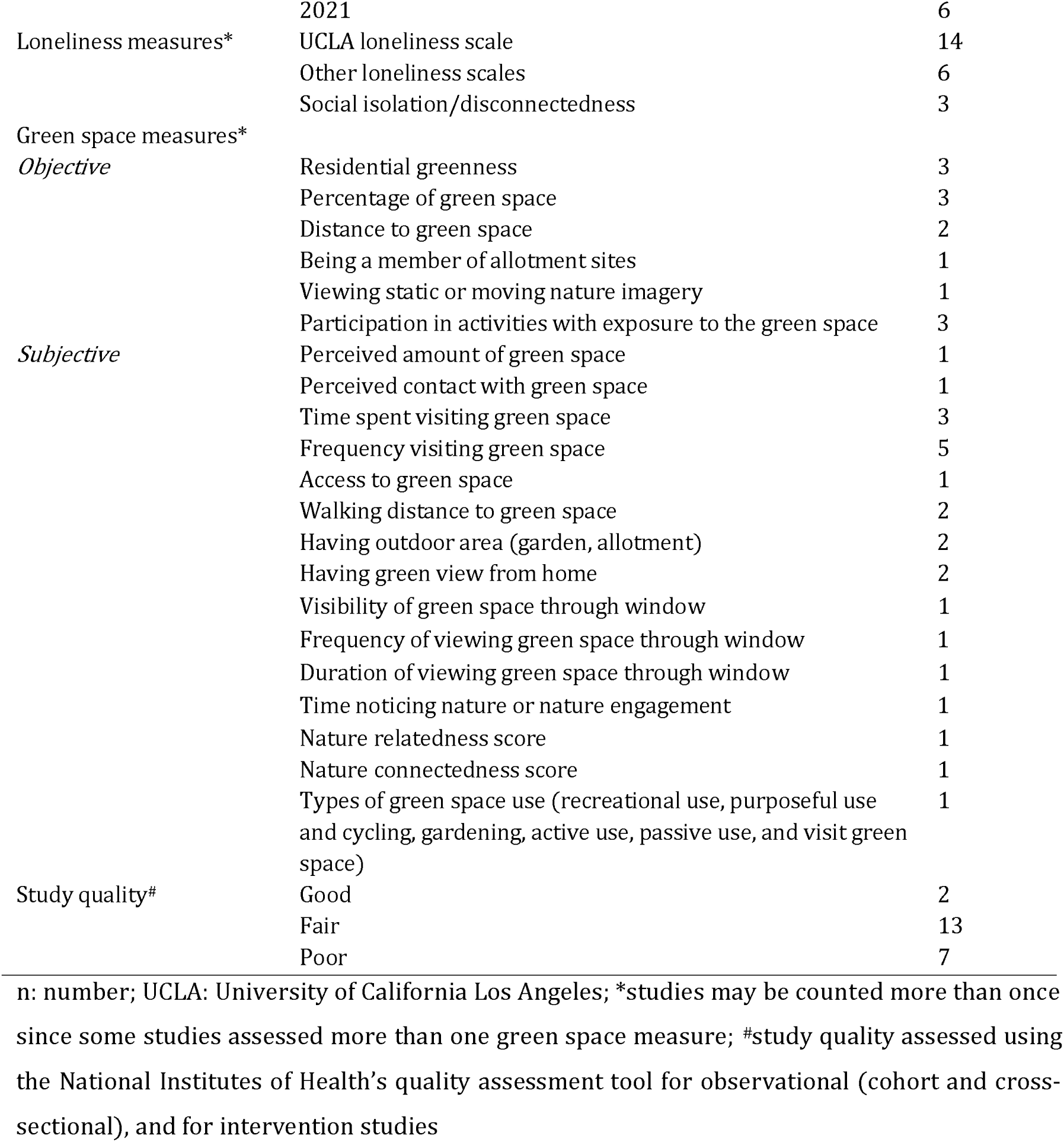
Summary of final studies reviewed

Each of the experimental studies had a small sample size (n<79) ^55 60 62 63 66^ and two cross-sectional studies analysed a sample of ≤200.^47 50^ The largest sample size was in a cross-sectional study in the UK with 209,525 participants,^54^ followed by a study in Germany with 17,602 participants.^56^ The two longitudinal studies had a sample size of 397 participants (11,193 assessements)^68^ and 8049 participants.^64^ While most studies were conducted among individuals ≥16 years of age, a few recruited older adults aged ≥50 and ≥60 years.^55 61 62 66^ One cross-sectional study had participants who were as young as 12 years of age.^49^ In addition, some studies only involved participants with specific characteristics, such nursing-home dwellers,^62^ registered elderly voters,^61^ and male prisoners.^65^

### Green space measures

Studies examined different subjective and/or objective measures of green space in relation to loneliness (Table 2 and Supplementary Table 1). Nine studies assessed subjective measures,^48 50 53 56 58 59 61 65 68^ three studies^52 57 67^ examined both objective and subjective measures, and the other ten studies used objective measures only.^47 49 51 54 55, 60 62–64 66^

The most common subjective measures were time spent visiting green space (three studies^48 57 58^) and frequency of visiting green space (five studies^52 53 57 59 67^). Other subjective measures included the perceived amount of green space,^57^ perceived contact with nature,^68^ having access to green space,^61^ walking distance to green space,^56 59^ having an outdoor area (garden, allotment),^52 59^ having a green view,^52 67^ visibility, frequency, and duration of viewing green space through window^65^, time noticing nature or nature engagement,^53^ and types of green space use.^50^ One study obtained a ‘nature relatedness score’ using the Nature Relatedness Scale^59^ and another a ’nature connectedness score’ using the Nature Connection Index,^53^ both taken to indicate the extent that participants felt connected to nature and/or natural settings, while not explicitly measuring contact with green space.

The objective measures such as percentage of green space or residential greenness within a particular buffer or administrative area were assessed using land use data or normalised difference vegetation index (NDVI) in six studies.^49 52 54 57 64 67^ Two studies objectively measured the distance to green space.^51 57^ One study assessed whether participants were members of allotment sites as a proxy for exposure to green space.^47^ Four studies used intervention-based exposure to green space that consisted of indoor gardening programs,^62 66^ a community intervention through visiting kitchen gardens and walking in neighbourhood green spaces,^55^ exposure to nature imagery in a lab setting,^63^ and a ‘park prescription’ which provided counselling about benefits of experiencing nature.^60^

### Loneliness and social isolation measures

The main data on loneliness came from self-report measures, wherein individuals were asked about time spent with other people at a specified time, how embedded they felt within groups of friends, how often they felt left-out and isolated from others, if they lacked companionship, as well as direct feelings of loneliness. With several exceptions,^47 51 53–55 64 68^ most studies used the UCLA loneliness scale,^69^ with variation between the three-item,^48 50 56 60 63^, six-item,^49 57 58 65^ and 20-item versions.^59 62 66 67^ Two studies examined social isolation^52 54^ in relation to green space, and another one assessed social disconnectedness,^61^ obtained using a single-item question. Loneliness or social isolation was assessed as a secondary outcome or as a candidate mediator in four studies.^49 52 57 58^

### Association between green space and loneliness

We extracted 132 associations from the 22 studies. These included associations for multiple measures of green space, and loneliness, as well as multiple associations stratified by different effect modifiers within individual studies (Table 3). The majority (n=88, 66.6%) were in the expected direction (negative): more green space exposure or experience was attended by less loneliness. Of the 88 associations in the expected direction, 44 (50%; or 33.3% from the total) were statistically significant (p<0.05). One study reported a statistically significant association in the unexpected direction.^53^

**Table 3.** Summary of associations extracted from 22 articles

| Green space masures | n<sup>a</sup> | Associations |  |  |  |  |
| --- | --- | --- | --- | --- | --- | --- |
|  |  | Significant<sup>b</sup> |  | Non-significant<sup>c</sup> |  |  |
|  |  | E<sup>d</sup> | UE<sup>e</sup> | E<sup>d</sup> | UE<sup>e</sup> | NR<sup>f</sup> |
| Objective measures |  |  |  |  |  |  |
| Residential greenness within buffers of buffers of: |  |  |  |  |  |  |
| - 100 m | 1 |  |  |  |  | 1 |
| - 250 m | 1 |  |  |  | 1 |  |
| - 300 m | 1 |  |  |  |  | 1 |
| - 500 m | 3 | 1 |  | 1 |  | 1 |
| Percentage of green space within buffers of |  |  |  |  |  |  |
| - within an administrative area | 1 |  |  | 1 |  |  |
| - 400 m | 2 |  |  | 1 | 1 |  |
| - 800 m | 2 |  |  | 2 |  |  |
| - 1 km | 17 | 6 |  | 8 | 3 |  |
| - 1.6 km | 33 | 7 |  | 14 | 12 |  |
| - 3 km | 17 | 9 |  | 6 | 2 |  |
| Distance to green space | 2 |  |  | 1 | 1 |  |
| Being a member of allotment sites | 2 | 1 |  |  | 1 |  |
| Viewing static or moving nature imagery | 8 | 2 |  | 3 | 3 |  |
| Participation in community intervention with exposure to green spaces<sup>g</sup> | 2 | 2 |  |  |  |  |
| Participation in park prescription group<sup>g</sup> | 5 | 5 |  |  |  |  |
| Participation in indoor gardening<sup>g</sup> | 4 | 2 |  |  |  | 2 |
| *Sub-total* | 101 | 35 | 0 | 37 | 24 | 5 |
| Subjective measures |  |  |  |  |  |  |
| Perceived amount of green space | 1 |  |  |  |  | 1 |
| Perceived contact with green space | 1 | 1 |  |  |  |  |
| Time spent visiting green space | 3 | 2 |  |  |  | 1 |
| Frequency visiting green space | 6 | 1 |  | 2 | 1 | 2 |
| Access to well-maintained green space | 1 |  |  |  | 1 |  |
| Walking distance to green space, sports, leisure facilities | 3 | 3 |  |  |  |  |
| Having outdoor area (garden, allotment) | 2 | 1 |  |  | 1 |  |
| Having green space view from home | 2 | 1 |  |  |  | 1 |
| Visibility of green space through window | 1 |  |  | 1 |  |  |
| Frequency of viewing green space through window | 1 |  |  | 1 |  |  |
| Duration of viewing green space through window | 1 |  |  |  | 1 |  |
| Time noticing nature or nature engagement | 1 |  | 1 |  |  |  |
| Nature relatedness score | 1 |  |  | 1 |  |  |
| Nature connectedness score | 1 |  |  | 1 |  |  |
| Types of green space use |  |  |  |  |  |  |
| - Recreational use | 1 |  |  |  |  | 1 |
| - Purposeful use and cycling | 1 |  |  |  |  | 1 |
| - Gardening | 1 |  |  |  |  | 1 |
| - Active use | 1 |  |  |  |  | 1 |
| - Passive use | 1 |  |  | 1 |  |  |
| - Visit green space | 1 |  |  |  |  | 1 |
| ***Sub-total*** | **31** | **9** | **1** | **7** | **4** | **10** |
| **Total: n (%)** | **132** | **44**<br>**(33.3)** | **1**<br>**(0.8)** | **44**<br>**(33.3)** | **28**<br>**(21.2)** | **15**<br>**(11.4)** |

<sup>a</sup>number of associations of between green space and loneliness that count multiple indicators of green space and multiple associations within a single study (e.g., analysis stratified by effect modifiers); <sup>b</sup>statistically significant association ( $p < 0.05$ ); <sup>c</sup>non-statistically significant association ( $p \geq 0.05$ ); <sup>d</sup>association in expected direction; <sup>e</sup>association in unexpected direction; <sup>f</sup>association in non-reported direction; <sup>g</sup>in-person observation used as data source for exposure to green space in experimental studies

#### Evidence from the longitudinal studies

Astell Burt et al’s^64^ study in Australia found a lower cumulative incident of loneliness (over 4 years) with an increase in urban greening within 1.6 km. This association was stronger in individuals living alone. Associations between green space within shorter distances (400m, 800m) and loneliness were weaker. A study by Hammoud, et al.^68^ involving 11,193 ecological momentary assessments nested within 397 participants indicated that contact with nature was associated with lower odds of loneliness.

#### Evidence from cross-sectional studies

Studies are presented by the type of green space measure analysed. Zijlema, et al. ^57^ assessed residential green space quantity within buffers of 100 m, 300 m, and 500 m in multiple cities. Soga, et al. ^67^ used a buffer of 250 m for measuring green space quantity. Neither study found a reliable association between objectively-measured green space and loneliness. Meanwhile, a study by Lai, et al. ^54^ with more than 200,000 participants found a statistically significant association between residential greenness within a buffer of 500 m and social isolation, but not loneliness. A study by Ward Thompson, et al. ^52^ found a small non-statistically significant association between the percentage of green space within an administrative area and social isolation. By contrast, findings from a study by Maas, et al. ^49^ in the Netherlands indicate that higher percentages of green space within 1 and 3 km radii were associated with lower odds of feeling lonely. That study also investigated modifying effects of age groups, education, household income, and urbanicity. Statistically significant associations in the expected direction were found among children, adults, and elderly, but not among youth, those with lower education, those with low household income, or those living in urban municipalities. Another study from the Netherlands^51^ and a multi-city study^57^ (comprising Barcelona, Spain; Doetinchem, the Netherlands; and Stoke-on-Trent, United Kingdom) tested and found no clear evidence of association between objectively measured distance to green space and loneliness. In addition, van den Berg, et al. ^47^ found age group moderated the association with loneliness of being an allotment gardener (as established by the researchers). Among participants 62 years and above (but not in other age groups), those with an allotment garden reported less loneliness than neighbours without one.

Perceived quantity of green space was not associated with loneliness in the multi-city study by Zijlema et al.^57^ This study reported no statistically significant association between residential distance (measured objectively as the Euclidean distance) to the nearest natural outdoor environment and loneliness,^57^ but another found lower levels of loneliness with more self-reported time spent visiting green space.^58^ Buecker, et al. ^56^ and van Houwelingen-Snippe, et al. ^59^ found that participants who reported longer walking distances to nearby nature, public parks, and sports and leisure facilities had a higher level of loneliness. Time spent sightseeing and visiting an amusement park and zoo was found to be associated with lower levels of loneliness in a Dutch study by MacDonald, et al. ^48^

Five studies tested for association between frequency of visiting green space and loneliness or social isolation.^52 53 57 59 67^ Only a study by Soga, et al. ^67^ conducted during the Covid-19 pandemic in Japan reported higher frequencies of visiting green space were associated with lower levels of loneliness. In addition, that study also indicated that having a green view through a window also potentially reduced feelings of loneliness. However, the study by Ward Thompson, et al. ^52^ using a smaller sample size and correlation analysis without control for potential confounders did not report an equivalent association between having a view to green space or hills with social isolation. Similary, a study among male prisoners by Li, et al.,^65^ found no associations between visibility, frequency, and duration of viewing green space through window and loneliness. A study of older registered adult voters in the USA found no association between having access to well-maintained and safe parks within walking distance and social disconnectedness.^61^

Two studies estimated the association between reports on having an outdoor area such as a garden or allotment with loneliness or social isolation.^52 59^ While no association was reported by van Houwelingen-Snippe, et al. ^59^, Ward Thompson, et al. ^52^ reported a statistically significant negative correlation between having access to an allotment or garden and loneliness, but without adjustment for possible confounders. In addition, a study by Bergefurt et al. showed that individuals who frequently used public space for passive activities such as sitting, watching and gathering were less likely to feel lonely, though the association was not statistically significant.^50^ The studies that obtained ‘nature relatedness’^59^ and ‘nature connectedness’^53^ scores did not find them statistically significantly associated with loneliness in the expected direction. A study by Richardson and Hamlin found a statistically significant association between time noticing nature or nature engagement with loneliness in the unexpected direction.^53^

Some of the cross-sectional studies assessed loneliness or social isolation as a mediator of the association between exposure to green space and health-related outcomes. Maas, et al. ^49^ found that loneliness mediated associations between percentages of greenness within buffers of 1 and 3 km and several health measures, including perceived general health, number of health complaints, and psychiatric morbidity. Similarly, van den Berg, et al. ^58^ reported mediation by loneliness of associations between time spent visiting green space and both mental health and vitality. Ward Thompson, et al. ^52^ indicate that social isolation mediated association between having an allotment or garden and perceived stress. However, no mediation by loneliness was reported by Zijlema, et al. ^57^ for association between distance to the nearest outdoor environment and cognitive function, possibly due to lack of clear association between the same green space exposure measure and loneliness.

#### Evidence from trial-based studies

Tse ^66^ conducted a quasi-experimental study of an 8-week indoor gardening program for nursing home residents in Hong Kong. The gardeners realized a significantly greater reduction in loneliness compared to controls. The results were corroborated by qualitative data which indicated some participants expressed less loneliness post-intervention. However, a similar study done in the US among older rural nursing home residents did not find a statistically significant difference in loneliness between those receiving a 5-week indoor gardening program and the control group that received a 20-minute visit during the same period.^62^ Both studies had small samples, used the UCLA loneliness scale, and had short intervention periods, but the latter^62^ lacked a proper control.

Razani et al.’s randomised trial with low-income parents found no difference in loneliness between those who received a park prescription only compared with those who also received additional enablers for park visits, indicating the enabling intervention did not have an extra effect on loneliness reduction at 1- and 3-month follow ups.^60^ Nevertheless, this study reported an overall reduction in loneliness in the whole group and a positive impact on park visits. Rodríguez-Romero, et al. ^55^ demonstrated that interventions comprising kitchen garden visits and walks through greener neighbourhoods as a part of a broader intervention package over 6 months did more to reduce loneliness than did care as usual. This study was conducted among persons >64 years old with some degree of lonely feelings and limited autonomy.^55^ Laboratory-based studies by Neale, et al. ^63^ found a reduction in loneliness scores among participants in a group with exposure to ‘nature’ vs. ‘urban’ imagery. There were no differences in loneliness scores between those exposed to natural or urban stimuli ‘with’ and ‘without’ people in the imagery shown.

## DISCUSSION

### Main findings

The balance of evidence indicates more green space is inversely associated with loneliness, with 88 of 132 (66.6%) associations reported in the expected direction and 44 (33.3%) achieving statistical significance (p<0.05). However, the quantity of evidence is currently low, with just 22 studies overall, of which most had only fair quality. The evidence is based mostly on cross-sectional data; there are few trials,^55 60 62 63 66^ and longitudinal studies^64 68^ are especially scarce. With two exceptions,^50 61^ the current literature is agnostic with respect to assessment of the different types and qualities of green space, and only two studies have considered whether loneliness mediated associations between green space and distal health outcomes. Only one study has examined a potential pathway linking green space with loneliness (via nature identity).^63^ Few studies have assessed effect modifiers, and these focused on individual differences (e.g. age,^47^ relationship status^64^). Contextual contingencies and different types of loneliness were not examined, nor were ecological studies conducted.

### Strengths and limitations

To our knowledge, this is the first systematic review providing a synthesis of current evidence on the association between green space and loneliness. We used PRISMA guidelines in developing and reporting the systematic review. Screening for eligible studies used seven frequently accessed databases, adopting keywords from previous systematic reviews, and checks on references of included studies bolstered comprehensiveness.

There are some limitations of the methodological aspect of this review and eligible articles reviewed. With regard to the review method, articles published in non-English are not included, and articles that deal with related concepts, such as social connectedness and social support, were not included, reflecting our reluctance to interpret low levels of such concepts as necessarily indicative of loneliness. With regard to the evidence reviewed, synthesis of findings indicates that, at this stage in the development of the literature, the evidence for association between green space and loneliness is weak; most of the studies included were cross sectional in design and do not support strong causal inferences. Different measures of green space yielded mixed findings on the association between green space and loneliness. Consequently, more studies with stronger designs are warranted to confidently make recommendations regarding the amount of neighbourhood green space needed; provisions for particularly important aspects of green space; and the design of interventions. Furthermore, most studies in this review were from high-income countries, and hence, the findings might not generalise to settings in middle-and low-income countries.^70 71^

### Theoretical and methodological guidance for future research

A more general limitation of the extant literature on green space and loneliness is the lack of a coherent, dedicated conceptual model integrated with wider research on nature and health.^21 23 26^ Such a model is necessary to guide future research that will better support practical applications.

As a starting point for the development of such a model here, we recognize that loneliness takes multiple forms and has diverse concomitants (as outlined in the Introduction). This recognition is required for elucidating the potentially multiple mechanisms that link experiences with different kinds of green space with different ways of feeling lonely, as well as specifying the circumstances upon which particular green space – loneliness associations are contingent.

Figure 2 fuses the results of this systematic review with findings from other relevant qualitative and quantitative studies to elaborate and extend a general conceptual model first proposed to clarify how contact with green space can lead to health benefits via multiple pathways, organized in domains defined in terms of their adaptive relevance (reducing harm, building capacities, restoring capacities).^21^ The model was recently modified to link health with biodiversity^26^ and wildlife,^72^ in each case expanded with the addition of a fourth domain of pathways, those by which aspects of biodiversity could cause harm. Innovations depicted by our conceptual model include: (a) acknowledgement of the potentially rich diversity of green space types, or green places to which a person may have access; (b) the level of congruence between personal and place-based differences in circumstances that condition a person’s susceptibility to both the benefits and dis-benefits of green space, and their capacities for engaging meaningfully with it; (c) the experiences (personal, relational, collective) a person may have with green space and and other people present or absent within it; (d) the domains of pathways through which green space experience engenders effects, customized to loneliness and its concomitants; and (e) the explicit positioning of loneliness and its concomitants as mediators within the broader causal system that links green space with health and wellbeing.^21 26^

**Figure 2:**
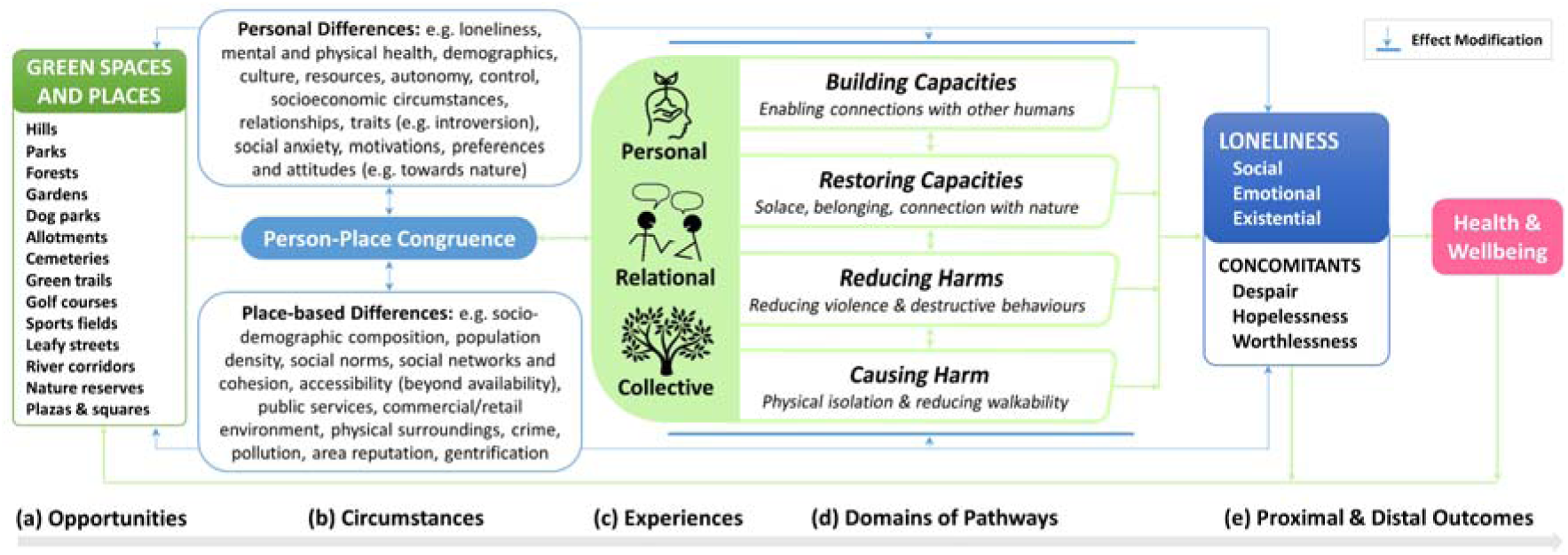
Conceptual model linking green space with loneliness and concomitants

We also acknowledge a complex circularity inherent to this system with arrows running bidirectionally between opportunities and experience, through circumstances, reflecting the understanding that, over time, a person’s or group’s history of experience with a given green space will feed back to shape the character of the opportunity recognized in the green space. Similarly influential feedback is represented by the arrows emerging from loneliness and health, running back to green space. In this regard, ample research indicates the propensity for relocating between neighbourhoods is highly selective and green space provision may be an important factor in choices made.^73^ Healthier people are more likely to move to less deprived areas^74^ which tend to have more green space (e.g.^75^), while people in poorer health may be either less likely to move home,^76^ or are more likely to relocate to affordable housing in deprived areas, given the documented relationship between health and socioeconomic circumstances.^77^ However, despite associations between loneliness and poor health being well-documented (e.g.^10^), the general understanding of loneliness as a signal for an individual to connect and satisfy some unmet need for companionship^78^ may drive people who feel lonely into areas with more green space in efforts to build or restore feelings of connection, unless they already feel socially isolated living in an area with few people but extensive green space.^79^ This potentially complex, bi-directional relationship between loneliness and residential mobility is under-researched and may have important implications for future epidemiological studies on the topic.

Hereafter, we discuss the major components of our conceptual model and in doing so, we offer guidance for future research designed to estimate association between green space, loneliness and its concomitants, as well as candidate mediators and potential effect modifiers.

#### (a) Opportunities

Inequities in the availability of green space^75 80 81^ generally and tree canopy cover^82–88^ in particular have been reported in various countries. This component of the model is described in terms of the existing ‘opportunities’ for experiencing green space, rather than ‘exposure’, to emphasise that (1) particular types of green spaces have been arranged, designed, and/or managed to serve particular sets of activities that serve particular sets of needs, and (2) they have acquired meanings over time that may also figure significantly in a person’s experience, on a given occasion and over repeated visits. Here we also want to acknowledge that the opportunities people have for experiencing green space may involve some degree of separation; a person need not be physically within a green space to appreciate, for example, the laughter of children playing there and other sounds that reach one’s window.^89 90^ This could even extend to simple knowledge of existence (e.g. through storytelling of historic events for maintaining cultural connections across generations^91^). This terminology is purposefully aligned with the ‘cumulative opportunities’ concept,^92^ in which the network of green spaces to which a person may have access is important, not simply the distance or travel-time to that which is nearest.

Through this model we also recognise that opportunities permitted and promoted by green spaces may be multifactorial within the same setting and distinctively clustered across settings. This acknowledges that the types and qualities of green space people can readily access are likely to be pivotal for reducing loneliness, with the ‘qualities’ aligned with the ways in which the given green spaces enable people to do things they think will enrich their lives.^93^ These qualitative differences, together with the meanings assigned to those green spaces over time, encourage a distinction between spaces and places, often applied in environmental psychology, human geography, landscape architecture and other fields. In the following, we will maintain the connection to the broader literature by using the term ‘green space’; however, it will become apparent that we are often referring to ‘green places,’ and that the distinction between space and place is relevant for some pathways to loneliness.

Take for instance two distinctive types of green spaces: cemeteries, which support remembrance of past lives, and allotments, which encourage nurturing of new life. It was notable that several of the quantitative studies we reviewed considered allotments^47 52 59^ and gardens,^55^ while none examined the role of cemeteries. However, qualitative research indicates both of these types of green spaces bring people together to bond over public rituals and physical activities.^94–96^ As settings where people visit, linger and interact, sometimes over many generations, these particular green spaces have been invested with particular shared meanings that can support and sustain cohesion and prosocial behavior in local communities.^39 97^ Were either of these types of green spaces threatened or neglected, those who have some relationship with them could be expected to act for their protection and maintenance.^98 99^ Should their decline be allowed to continue, their potency for generating and strengthening connections between people would likely be vastly diminished and may even signal a community in decline.^100^ Yet, along with these commonalities, the two types of green place also serve particular functions, and so have special values and involve behavioral norms and management practices with a bearing on relief (or aggravation) of particular forms of loneliness. For example, a common scenario in popular literature and films involves a person having a graveside conversation with a lost loved one, the absence of whom is profoundly painful and unsettling, and continuing visits to whom offer comfort and stability in the grieving process.^101^

Future work needs to theorise how different types and qualities of green spaces and places may be connected, for good or ill, with specific types of loneliness and its concomitants, and proceed to measure and map inequities in the cumulative opportunities available. To this point we have focused on green space opportunities that currently exist for a person. Yet, diverse contextual factors determine which green spaces and places are available. For example, substantial literature has documented inequities in the availability of green space^75 80 81^ generally and tree canopy cover^82–88^ in particular, within and across varous countries. In addition to closer attention to the significance of activity affordances, behavioral norms, and meanings particular to different types of green spaces, future work needs to measure and map the direct effects of inequities and other contextual variables on the availability of opportunities. This work will complement efforts to understand how the circumstances of people who could use a green space shape the ways in which they engage with and experience it as well as the pathways from their experiences to proximal and distal outcomes. We turn now to consider those circumstances.

#### (b) Circumstances

Relations between green space, loneliness and its concomitants are likely to be sensitive to a complex interplay among personal and place-based differences in circumstances. These circumstances have import right the way across Figure 2 beyond traditional conceptualisations of effect modification, from determining opportunities for contact with green space, through to modifying the potency of various pathways and so net-impacts on loneliness. Here we reflect on key stages of influence.

##### Shaping opportunities for contact with green space through multilevel processes

Circumstances can directly affect opportunities for contact with green space by influencing their availability, while also shaping the risk of becoming lonely through impacts of demographic and socioeconomic change on the local environment, including the quality and quantity of green space and the social characteristics and activities of people in the green space. A notable and well-known example occurs at the level of the individual person, with lower personal socioeconomic circumstances usually restricting choice to more affordable housing stock often located in poorer neighbourhoods.^102^ These are typically less expensive in part because they have lower quantity and/or quality green space.^103–105^ Conversely, individuals with higher incomes and other advantages are able to exercise preferences with greater options in the housing market by purchasing property with or near to things that nourish their lives, like more and better quality green spaces (and in many contexts, blue spaces such as coastal and beach communities). Over time, the accumulation of these individually selective migrations is known to aggravate geographical segregation between communities by various demographic and socioeconomic characteristics.^106 107^ These population-level migratory processes also concentrate fiscal and political power that can help affluent communities preserve and maintain local green spaces, with those elsewhere left more vulnerable to dilapidation and elimination via “redevelopment”.^108^ The net result is not only a widening of spatial differences in health and wellbeing,^109^ but also the perpetuation of inequities in the quantity and quality of green space available, with flow-on effects for the risks of loneliness and its concomitants for current and future residents. Thus, while there is a tendency for research to be done with data at the level of the individual, this suggests that there also remains a need for investigation of how migration flows and selective (im)mobility (including aspects of gentrification and displacement, which we turn to later in a section on potential harms) influence geographies of loneliness and associations with green space availability using area-based longitudinal data analyses.

##### Person-place congruence in circumstances

An intermediate and hitherto under-recognised next step in the sequence depicted by Figure 2 is the interface of multilevel circumstances that we refer to as “person-place congruence.” This is an explicit recognition that the degree of alignment between personal and place-based circumstances can shape if and how people interact with nearby green space and the degree of susceptibility an individual exhibits towards it. Therefore person-place congruence, or lack thereof, has potential to unleash or mute specific domains of pathways linking green space with loneliness and its concomitants, before levels of magnitude are estimated. Thus, incongruence in these circumstances has potential to sabotage potential positive influences of green space on loneliness, or exacerbate negative influences, in complex ways. Qualitative and some quantitative studies provide rich illustration of the importance of person-place congruence and underline why it is important to consider both personal and place-based circumstances simultaneously in future research. Here, we present a suite of examples with emphasis on place and personal circumstances, while also providing some reflections on intersecting issues of temporality and lifecourse.

Many place-based sources of incongruence are remarkably common, despite their negative impacts being relatively well-known or self-evident. For example, typical features of cities such as major roads with inadequate crossing infrastructure, dilapidated footpaths and scarcely inclusive alternatives to steps and stairways can present significant barriers to visiting green space in general, and especially for people living with disability.^110–112^ Such circumstances can spatially marginalize and entrench feelings of being “out of place” among people with disability,^113^ who are already vulnerable to loneliness^114 115^ and therefore have high potential to benefit from the enrichment of environments to support social connection.^116^ However, place-based sources of incongruence can also emerge from efforts to be inclusive. For example, permitting of dogs to be off-leash will attract dog owners to green space and support associated benefits (e.g. walking^117^), but this can also discourage visits by those who worry about aggressive dogs and associated incivilities.^118 119^ Finally, some sources of place-based incongruence can stem from actions to intentionally exclude. Examples include the replacement of simple park benches in many cities with ones designed to prevent homeless people from sleeping on them (e.g. curved, hard surfaces).^120–122^ Even more overt are the rising levels of surveillance and privatisation of green space, both temporarily for commercial activities and also entirely with large areas fully under the jurisdiction of corporations, that can foster unpleasant feelings of being monitored and signal that certain groups of people and particular activities are not welcome.^123 124^

Emphasis on personal-differences in circumstances is also warranted and may further help to explain heterogeneity in prior results for green space, loneliness and health more generally. For instance, emerging research highlights adolescents higher in introversion and/or neuroticism personality traits, who have increased risks of loneliness,^125^ tend to benefit more from having quality green space nearby than their more extraverted and/or emotionally stable peers.^126^ Conceivably, these personality traits increase an individual’s susceptibility to stressful antecedent conditions and therefore create differentially greater potential for relief from social anxieties and chronic rumination through processes of restoration and social (re)connection promoted by green space.^43^

Another potential case of differential susceptibility involves the extent to which people seek contact with green space because of intrinsically motivating reasons to do with personal interest and emotional connection (perhaps aligned with “nature connectedness” and “nature relatedness” concepts), or extrinsic factors such as peer pressure, felt risk of social alienation, or some form of economic incentive. Self determination theory proponents^127^ have evidenced how behavioural change can be sustained through leveraging intrinsic motivations, whereas the effectiveness of strategies that apply extrinsic motivational techniques are not only shortlived, but may also undermine intrinsic motivations. The implications of this theory are that the utility of nearby green space as a passive intervention for reducing loneliness and its concomitants may be more effective in individuals with high levels of intrinsic motivation for engaging with nature, but the use of interventions that employ extrinsic techniques (e.g. a “nature prescription” from a health professional) may in some circumstances have unintended consequences.

Similarly, expectations have important roles in framing the extent that nearby green spaces are considered as attractive opportunities for experiences that could engage restoration or other pathways. For example, some of the people interviewed by Rupprecht, et al^128^ felt that informal green spaces have authenticity that is lacking in other types: *“It’s real, not fake like a park.”* This may extend to whether the quality of a green space matches how a person thinks it ought to be, whether in some ideal world or as it was in some remembered past. This is illustrated by Birch et al^37^: *“If you… walk through like parks and you look at the playgrounds that haven’t been done up in twenty years, and everything’s falling apart, it makes some places that should be happy more miserable.”* This scenario bears distinct similarities to the concept of ‘solastalgia’ introduced by Albrecht and colleagues,^129^ in which the observable degradation of environmental systems is considered to induce psychological distress akin to grief (also see^130^). In such circumstances, it may be that having dilapidated green spaces nearby aggravates, rather than provides relief from, loneliness and its concomitants, reminding of times and people past and gone.

Importantly, many of the aforementioned personal and place-based circumstances are subject to change over time, and may also be subject to conditioning based upon prior experiences (e.g. in early life). Some are characteristics of people, such as age-related ability, or of relationships between people, as with the formation, deepening, or dissolution of intimate relationships. Others reflect adaptations people make to local circumstances to secure benefits from green space, as with changes in working hours or commuting modes to enable more frequent visitation. Still others are characteristic of the broader social and cultural context; changes in these variables may simultaneously affect characteristics of green spaces, the people who can experience them, and the experiences they might have with them. These include ongoing urbanization, changes in occupations and lifestyles, and what many see as a widening disconnect between people and the natural world.^23 108^ It is therefore realistic to anticipate that pathways from green space to loneliness are subject to effect modification by variables at multiple levels over different stages of the lifecourse.^79^ Understanding of the relationships in question will gain from studies that are explicitly multilevel, taking into account person-place congruence. For example, scenario-based experimentation has illustrated how appreciation for a single possible visit – walking in a forest^131^ or sitting in a park^132^ – depends upon both current levels of cognitive fatigue (i.e., a need for restoration that varies within a person across time) and whether the person would be alone or in the company of a friend (also see ^42^). Understanding should also improve with studies that apply longitudinal designs, as exemplified by the one longitudinal cohort in our review, which could identify determinants of the incidence of loneliness, including green space opportunities which a person could have taken advantage of over a period of years.^64^

#### (c) Experiences

We assume that pathways from green space to loneliness run through experience. The presentations of the established^21^ and recently extended^26, 70^ pathway domain models have primarily focused on individual-level processes and the consequences of personal experiences with green space. We see a need to consider experience on additional levels of analysis, with a particular view to experiences shaped and shared by multiple individuals.^133^

Recognition of this need is exemplified by recent theorizing concerned with pathways in the restoration domain. Restoration processes emphasized by the extant pathway domain models have focused on ways in which ongoing adaptation to everyday demands drains psychophysiological and cognitive resources that individuals need to mobilize and direct action. These resources can be replenished through experiences with green space, according to attention restoration theory (ART)^134^ and stress reduction theory (SRT; also known as psycho-evolutionary theory).^135^ Applications of these theories in the green space and health literature, including that on loneliness synthesized in this review, have been largely agnostic to other scales at which experiences with green spaces and places can carry restorative processes that may be of equal, if not greater relevance to the outcomes of interest. This includes the experiences of green space that manifest on the scales of relationships between individuals in small groups (e.g. couples’) and larger collectives (e.g. communities), which may be crucial to expanding knowledge and informing potential policy options.

Two recent theoretical additions call attention to restorative processes that work on these higher levels of analysis. Relational restoration theory (RRT)^43^ emphasizes the extent to which experiences with green space can permit and promote pro-social interactions and supportive exchanges between individuals in close relationships. This can among other things restore relational resources should they have become depleted, which may in turn help to reduce loneliness. Collective restoration theory (CRT)^43^ refers to the attenuation of demands and promotion of positive shared experiences within local communities, cities and societies that may result from policies that enable widespread and simultaneous green space visitation and so the potential spread of benefits among those who come together in them, whether known or unknown to one another (e.g. public park provisions, public holidays, national vacation legislation^136^; see also ^38^).

Application of multilevel theorizing in future research is needed to understand the ways in which experiencing green spaces may not only help to permit and generate meaningful relationships that alleviate and prevent loneliness, but also the extent to which loneliness is reduced via individual and shared processes catalyzed by individual actions and exogenous factors that shift how entire communities view, relate to and interact with green spaces. These multiple scales of experiences with green space and the personal, relational and collective processes aligned with them are then manifest across all four domains pathways that extend towards loneliness and its concomitants.

#### (d) Domains of Pathways

Consideration of these multilevel processes of restoration has informed our integration of theories linking green space with loneliness and its concomitants within domains of pathways described in precursors to the present model.^21 23 26 70^ An important aspect of this integration is recognition of three possibilities: experiences in green space can engage multiple pathways simultaneously; these multiple pathways may be in the same domain (i.e., share the same kind of adaptive relevance) and/or in different domains; and multiple pathways may complement or compete with one another in the generation of effects.

The three domains of beneficial pathways (Building Capacities, Restoring Capacities, and Reducing Harm) from the original model,^21^ plus the fourth (Causing Harm) recently added,^26,70^are all retained in our conceptual model (Figure 2). As described in ‘circumstances’ and in keeping with an earlier conceptual model ^23^, pathways in all of the domains are subject to effect modification and, therefore, are candidates for moderated mediation analyses as well as mediation tests that address the ways in which mechanisms engaged along different pathways may work together or at odds with one another.^137^ .

##### Domain 1: Building capacities

Perhaps the most intuitive link between green space, loneliness and its concomitants involves building social connections. Whether they engender connections characterized as strong or weak^138^, pathways in this domain work to prevent loneliness. Green spaces can constitute pleasant, free-to-enter ‘Third Places’^139^ where people can go to satisfy momentary desires for more social interaction, thereby staving off more persistent feelings of loneliness.^140^ This may occur through serendipitous pro-social encounters, planned gatherings, and/or daily shared rituals.^43^

A fair amount of literature bearing on green space and health has addressed the workings of this kind of capacity building, for individuals and on the nighbourhood or community level^23^; however, relatively little has focused on loneliness as an outcome. The one longitudinal cohort study in our review indicated that those with more than 30% green space within 1600 m of the residence had less incident loneliness than those with less than 10% after four years of followup, while those who reported often feeling lonely at the start of the period did not report more relief at followup based on the amount of green space^64^. This speaks to prevention; however, as we will note later, this does not exclude the operation of restorative pathways, as the loneliness measure used did not differentiate among types of loneliness. The study also lacked relevant measures of social connection that could be used to directly test mediation hypotheses.

Qualitative evidence, however, indicates how green spaces can reinforce and foster new ties that evoke the warm feeling of embeddedness within community.^39 97^ Findings from semi-structured interviews of community garden members reveal how their participation is wrapped up in a sense of connection and camaradery.^94 95^ These green spaces may provide readily identifiable places where people can seek connection with others who share similar interests. They may also help to compensate for a lack of other green space in dense multi-family housing (e.g. apartments),^141^ which might contribute to loneliness. Similar might be observed of sports fields, which can serve as spaces not only for physical activity, but also for communual gatherings, cheering, marvel, bonding and formation of shared memories that can stimulate and reinforce a sense of belonging. This is likely facilitated by programming, which can activate green spaces as sites for volunteering and regular activities that engender feelings of belonging, as documented by interviews of participants on ‘Parkrun’, which operates in 23 countries around the world,^142–144^ for example: *“The real motivation for coming is the community thing. I always know that I’ll see someone I know and I nearly always end up talking to someone I’ve never met before”* (pp.97^144^).

##### Domain 2: Restoring capacities

Support for building social connections is the obvious, but not necessarily the only mechanism by which green space might reduce loneliness. We already indicated in the section on ‘experiences’ that restorative processes may work on multiple levels. Here we can elaborate on processes of restoring capacities as distinct from processes of building capacities. For instance, evidence by Maas, et al. ^49^ in this review indicated participants living in greener areas had lower odds of feeling lonely, while at the same time, those with more green space nearby did not experience more interactions with friends and neighbours, indicating alternative mechanisms to those aligned with the building capacities domain. The Restoring Capacities domain may involve one or more of at least three related mechanisms with green space supporting restorative experiences in solitude and providing relief for people experiencing the distress, distrust and lack of felt safety that characterises loneliness.^78^

Firstly, green spaces and community gardens in particular may serve as ‘affective sanctuaries’, permitting therapeutic settings for people experiencing emotional and physical exhaustion (i.e. ‘burnout’) to feel a sense of refuge. Or for those experiencing feelings of existential loneliness stemming from a sense of liminality (e.g. due to the diagnosis of a terminal illness), experiences with green space can afford opportunities for reflection on the meaning of these health states for an individual’s sense of self and hope for the future. ^145–147^ While this may be in the company of others, the restorative benefits may not necessarily require any direct interaction with other humans.

Secondly, regular momentary sharing of green spaces with other people, but without necessarily any direct interaction, may still generate a sense of undemanding connectedness and belonging to community. For example, interviews by Neal et al (2015, pp.472-473): *“In the park you feel like you’re kind of interacting even if you’re not speaking with them directly, but you’re sharing the space together […] you’ve both come to the park to enjoy what it is”.*^39^ This may be closely entwined with activities that result in the same people regularly visiting the same green spaces for the same, shared reasons, for instance, in the case of hillwalkers and those who walk dogs.^148^

Thirdly, ethnographic research indicates experiences with green space can evoke comforting memories that provide solace, which may be intentionally without the company of other humans.^40 41^ Interviews by Birch, et al. ^37^, for example, indicate that some people seek nature in solitude for its provision of non-judgemental, ego-free and dependable support: *“it’s just like the idea of being around nature I find very soothing. I think it’s ego free… nature doesn’t judge you.”*

This quote highlights what we call the ‘lean on green’ hypothesis, in which feelings of loneliness might be alleviated and/or prevented through establishing felt connection with the ‘more than human world’ and processes of restoration permitted and promoted by contact with nature, absent other humans. This might involve green spaces facilitating experiences with animals, such as dogs for which there is well-documented evidence of mental health benefits.^149 150^ Beyond pets, evidence indicates that visiting green spaces can afford sublime and life-affirming experiences that evoke wonder, awe, inspiration, and reverence for nature.^37 151 152^ Some studies report that a felt affinity for nature is associated with greater levels of eudemonic and hedonic wellbeing^153 154^ and pro-environmental behavior.^155 156^ Positive self-concept is purported to mediate reports of a so-called ‘warm glow’ following engagement in pro-environmental behavior^157^ (i.e. because a person feels it is the right thing to do or they are fulfilling a personal interest). Recent work indicates that a sense of meaning in life resulting from environmental engagement may also help to reduce loneliness.^158^ Mediation analyses are needed to quantify the contribution of the ‘lean on green’ hypothesis to association between green space and loneliness, from those which are likely to be supported by restoring social connections, provision of affective sanctuaries, and promoting a sense of belonging to community.

##### Domain 3: Reducing harms

Depression, despair, hopelessness, reckless risk taking and self-destructive behaviour are all concomitants of loneliness^35^ that, collectively, reflect on loss of meaning in life.^36^ Increasing evidence indicates that contact with green space may help to reduce these states, the antecedent conditions that sustain them, and the harms that can eventuate if action is not taken. Indeed, it may be that reducing harms is necessary for the effects of other domain pathways to flourish.

For example, qualitative research indicates violence in the community can result in people confining themselves indoors, inducing social isolation and potential loneliness.^159^ Greening may mitigate this harm. Evidence presented in a review by Mancus and Campbell (2019) concluded that *“the perception of safety is supported by quality, accessibility, and aesthetic dimensions of neighborhood [sic] green space”.*^160^ Another review, by Shepley, et al. ^161^, found that ca. 70% of the included studies (9 of 13) reported lower levels of crime in areas with more tree canopy cover (e.g.^162^). There is also an increasing number of pre-post intervention studies that report reductions in crime overall, and gun violence in particular, within communities where vacant areas of land have been cleared of refuse and grass and trees have been planted (e.g.^163 164^). Interestingly, these studies not only confirmed that residents in the intervention areas felt safer, but also reported improvements in mental health and more time outdoors spent relaxing and socialising.^164 165^ This further illustrates how pathways within different domains are interdependent and potentially synergistic in their operation.

There are many other examples emerging for the harm reduction domain pathway that have potential synergies with other pathways. For example, it is plausible that restorative processes at personal, relational and collective scales combine with lower levels of neighbourhood violence and increased social connection to facilitate reductions in pain,^166–168^ cigarette smoking,^169^ opioid dependence,^170^ substance misuse in adolescents,^171^ and risks of self-harm and suicidal ideation.^172–175^ Each of these emerging harm reduction pathways warrant further investigation, potentially with moderated mediation models and other methods that may be suitable to distinguish between pathways operating in serial or parallel.^137^

##### Domain 4: Causing harms

In efforts to understand the health benefits of green space, few studies hypothesise or test plausible ways in which urban greening may have direct or indirect unintended consequences that could result in harms to health.^176^ Yet, some studies^177–180^ have found higher risks of poor health, obesity and mortality in areas with more green space. A potential contributor to these counterintuitive results involves public green space that is low in quality and not attractive to visit, or that is private and unaccessible (e.g. private golf courses, agglomerations of large back gardens in suburban sprawl); such spaces may actually reduce walkability^181^ and opportunities to interact with neighbours.^49^ This may be aggravated further by perceptions of the behaviour of other people in those green spaces that discourage visitation, as was found in interviews by Byrne ^182^: *“I never go there because there are a lot of people drinking. I am afraid that they are going to do something to me…I don’t go because of the people.”*

Gentrification stands as an important example of how secular shifts in the local social context can have such multi-pronged effects along pathways that engender differing degrees of loneliness. ^183 184^ With gentrification, the scarcity of green space within a city is implicated in rising housing costs in greener neighbourhoods to levels that can be unaffordable to some residents, resulting in exclusion from their homes. Anguelovski, et al.^108^ indicate this can result in “social, cultural, and mental displacement, and dispossession,” with remaining residents losing neighbours and sense of belonging.^185, 186^ Thus, it may be possible that for some residents, the increasing availability of green space may come to be associated with higher risks of loneliness due to the loss of community belonging brought about by gentrification.

## Conclusions and future directions

This review reveals the quantum of evidence linking green space and loneliness remains small, limited mostly to studies of cross-sectional design, and absent of a clear conceptual model. We have provided such a model, together with theoretical and methodological guidance for future work. Our framework elaborates and extends existing frameworks,^21 26^ and is informed by findings from a range of qualitative and quantitative studies on related topics (e.g. despair, violence, gentrification, attitudes to nature). As the dire public health and societal consequences of inaction on loneliness and its concomitants continues to increase in the public and policymaker consciousness, it is clear that some well-meaning planners and health professionals will be motivated to ‘green’ our cities and support interventions to get people into green space (e.g. ‘nature prescriptions’), assuming benefits will come. That may eventually occur, but our review indicates a significant, persistent deficit in our knowledge of the potentially many ways in which experiences with green spaces influence loneliness and its concomitants. Overcoming these deficits should increase the linkielhood that interventions are viewed as credible in decision-making situations. There remains a chasm in our understanding of for whom the effects of green space might be sufficiently potent to bring about the desired results and for whom additional support is required. Finally, in the context of finite resources for preventive health strategies where there is already established evidence, for investing in green space and mechanisms to enable more time in green space for people who would benefit from it, we need studies to show how these types of interventions might be implemented effectively, cost-effectively, and sustainably and coordinated with efforts to address other major societal challenges, including climate change and biodiversity loss, without which ours may become a lonely planet indeed.

## Data Availability

All data produced in the present work are contained in the manuscript

## Acknowledgements

We thank Dr Tewodros Hailemariam for helping to conduct some of the early literature searches on a related topic.

---

**Supplementary Table 1.**
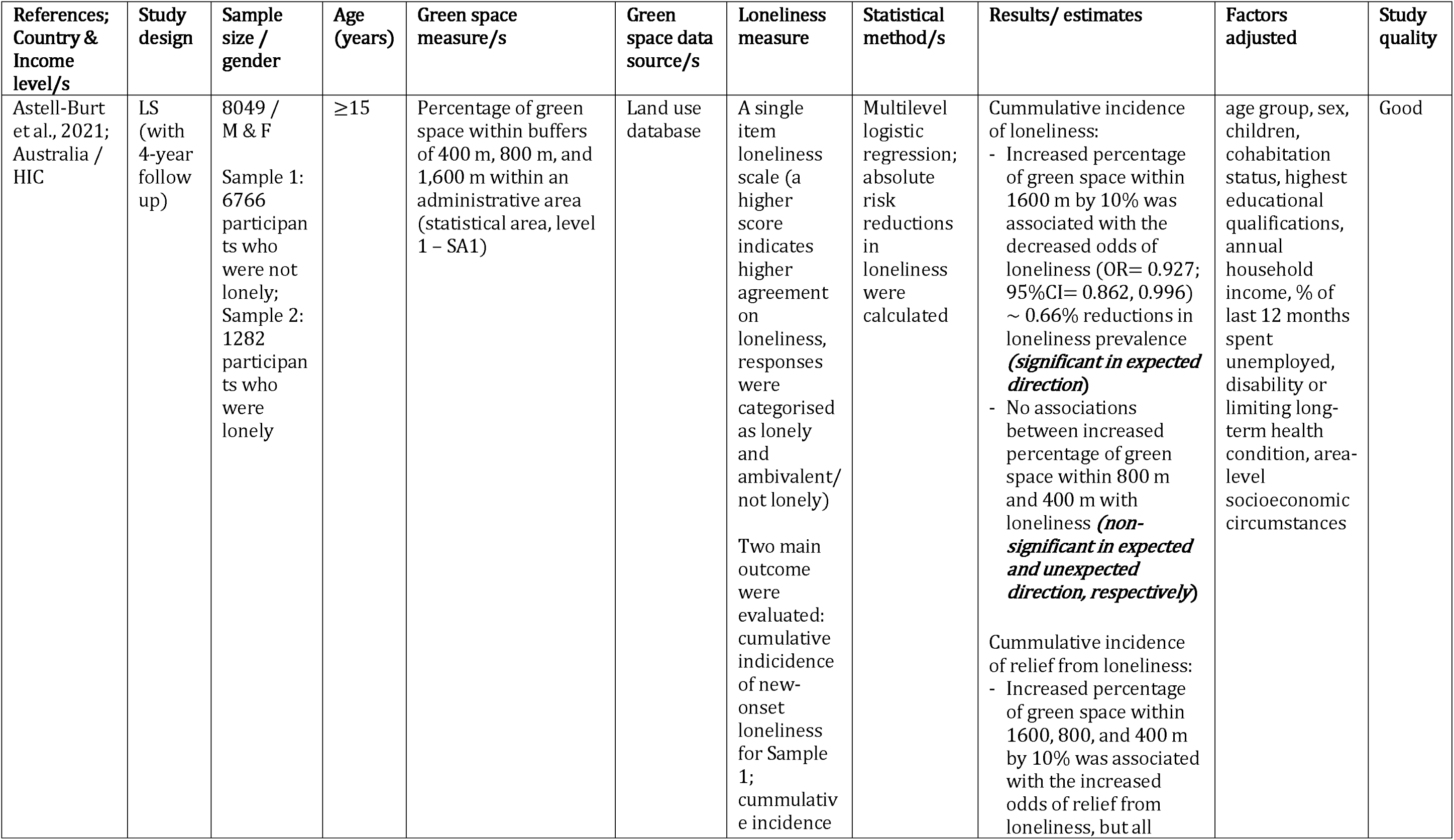
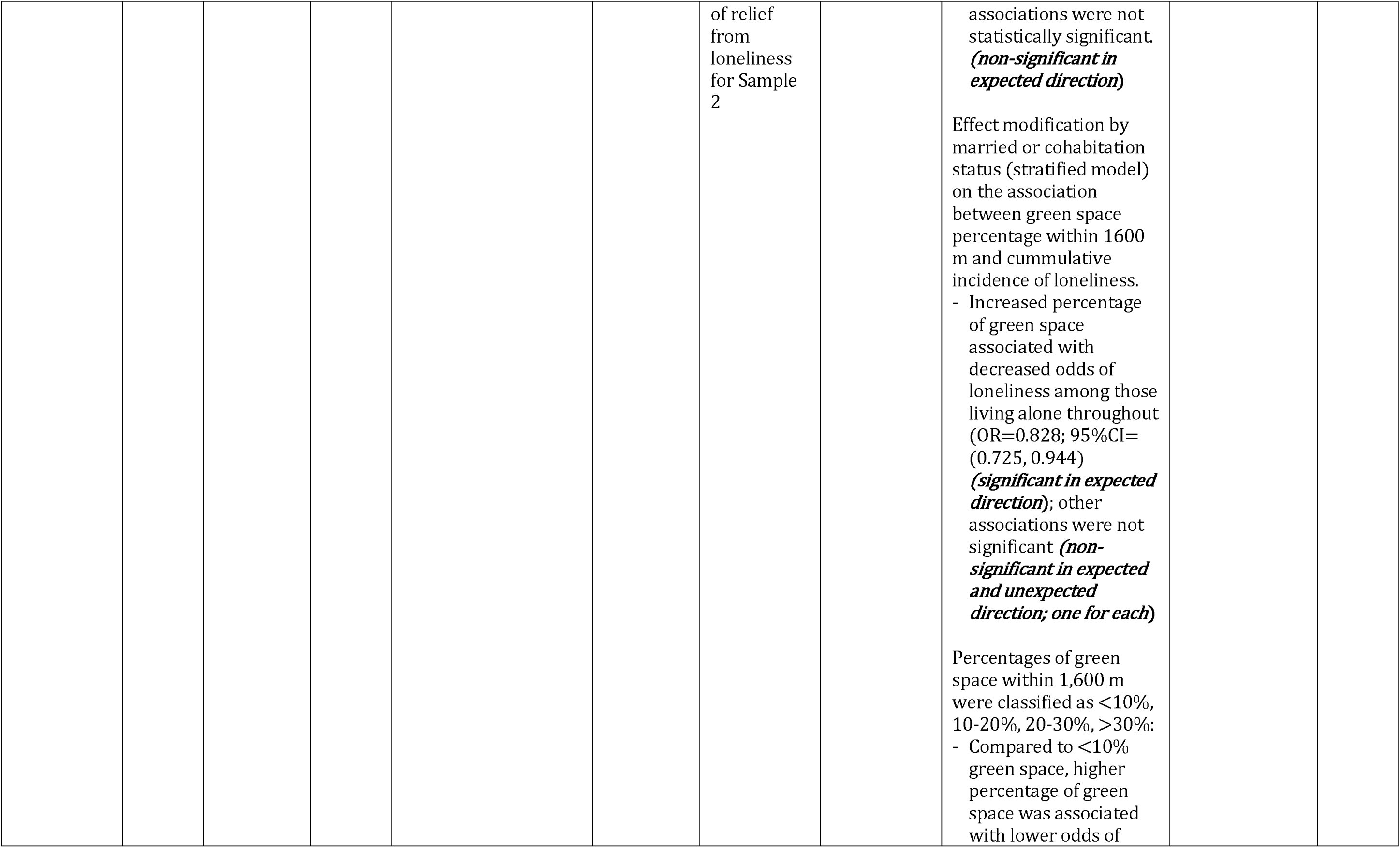
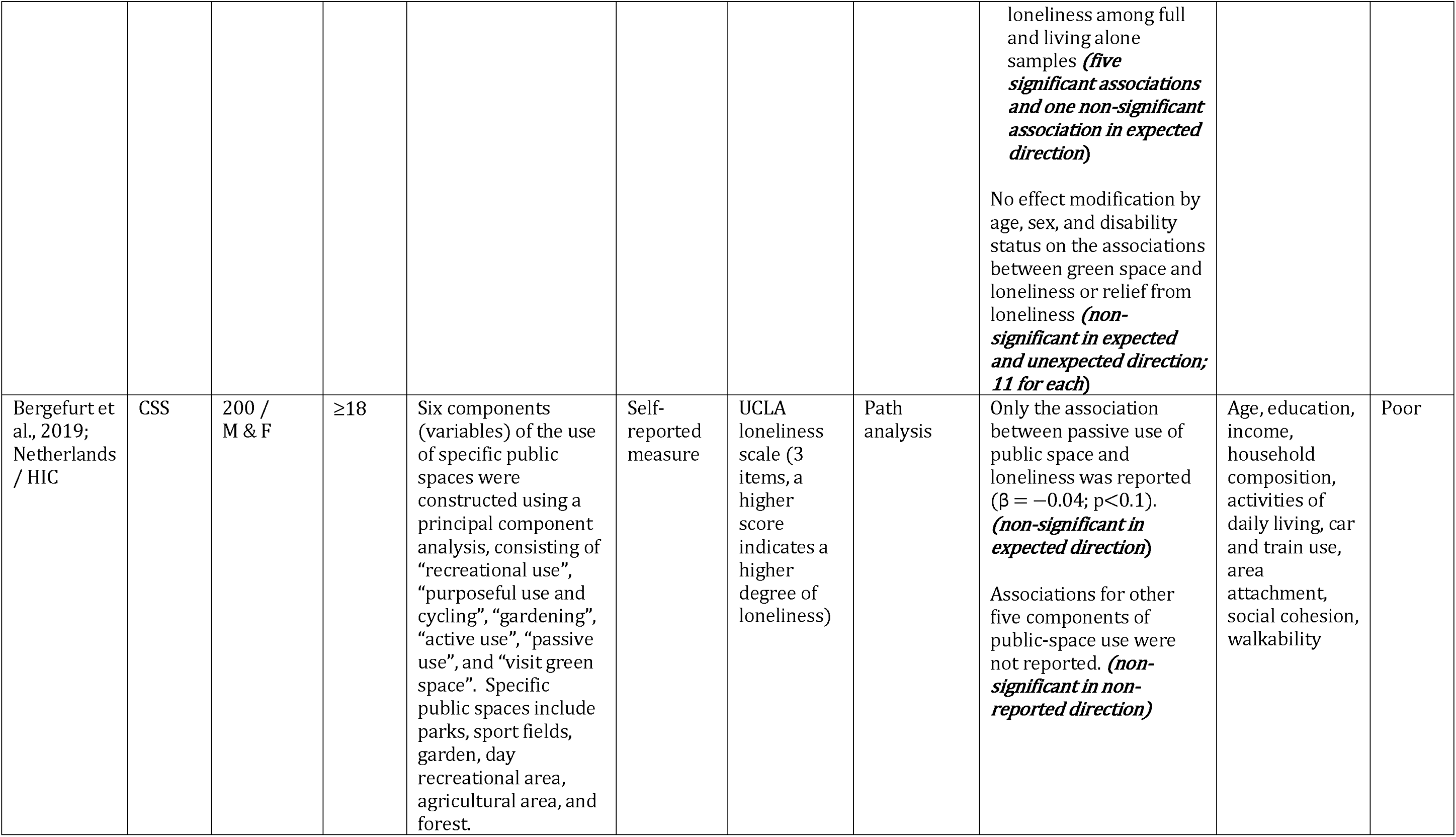
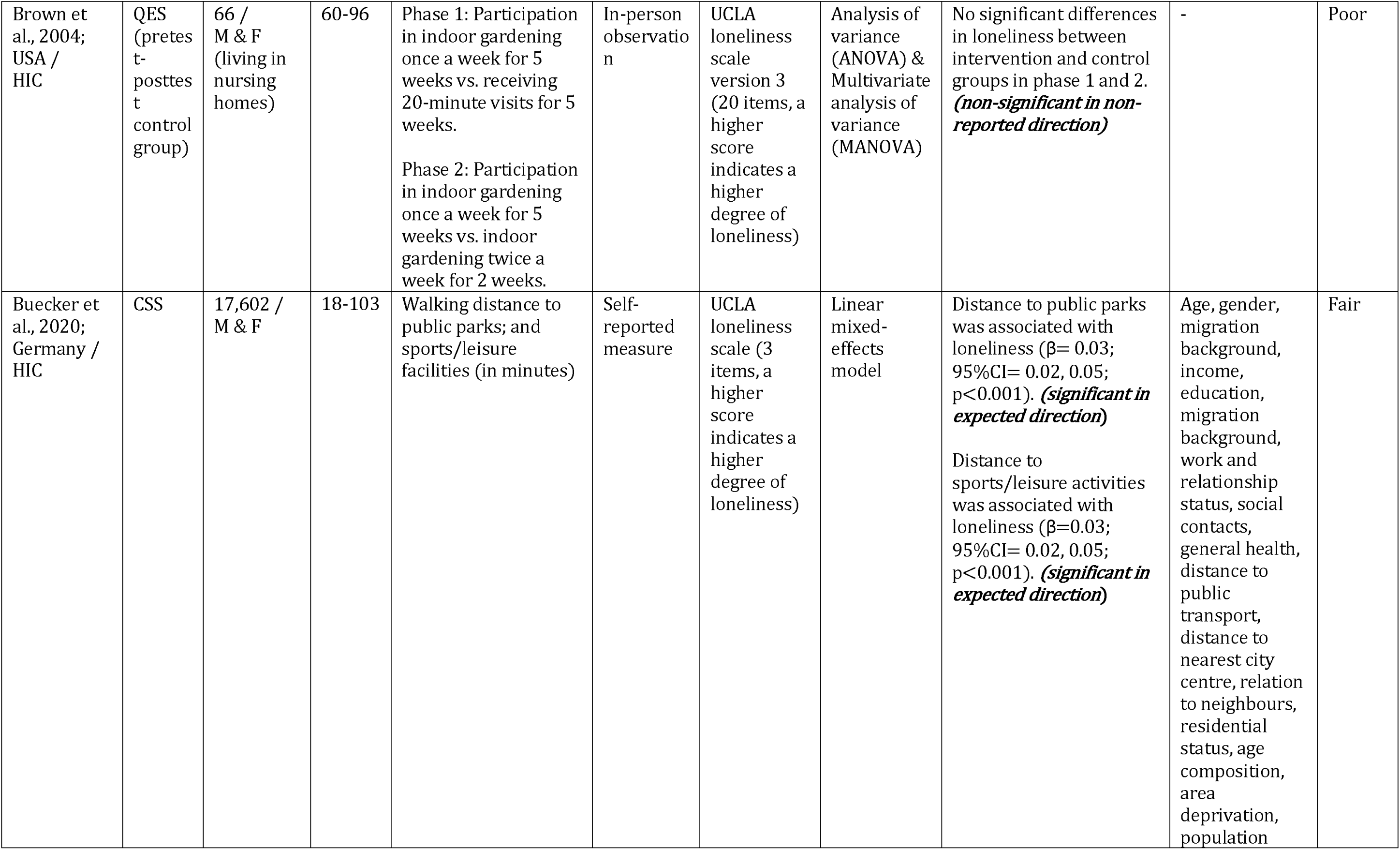
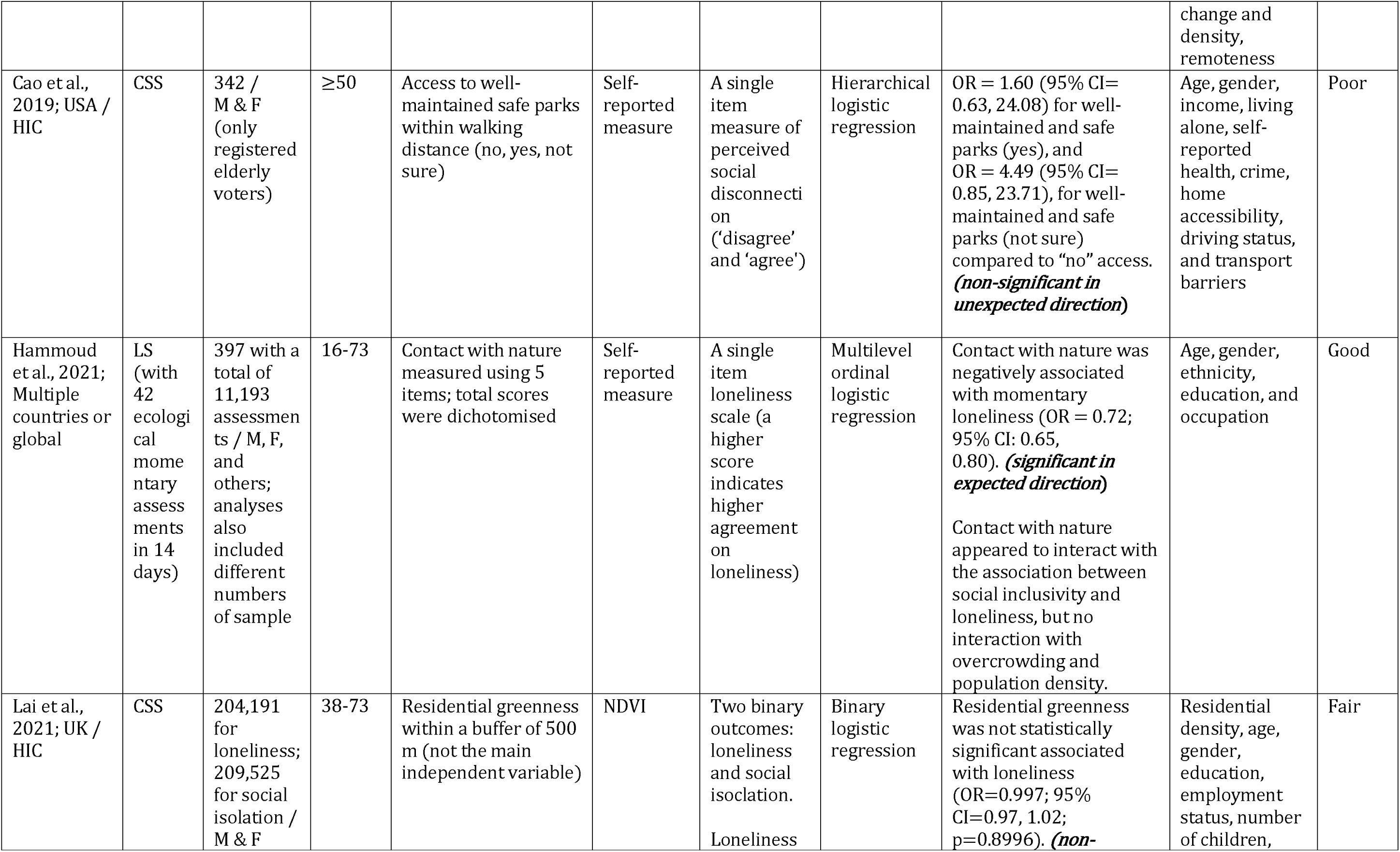
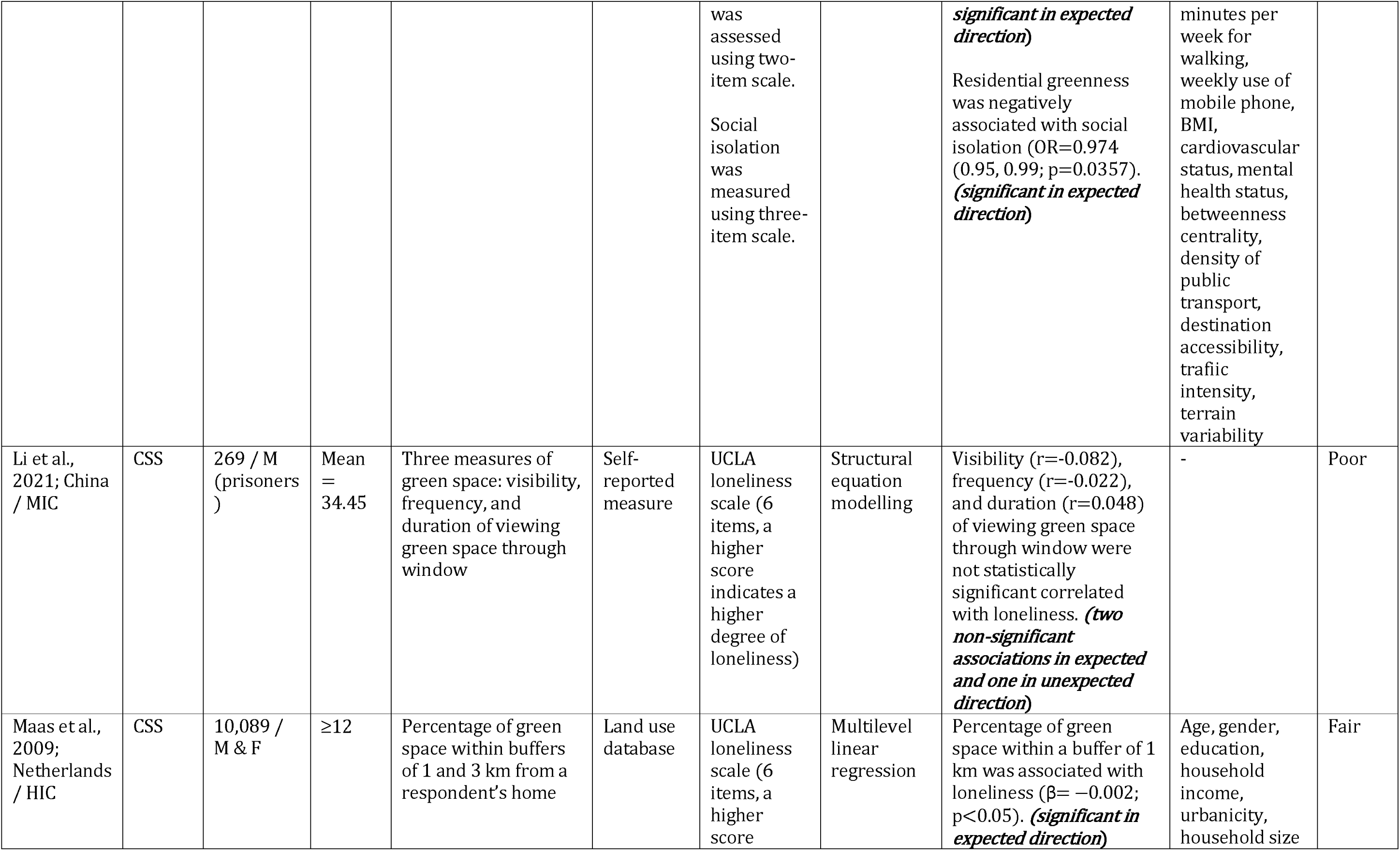
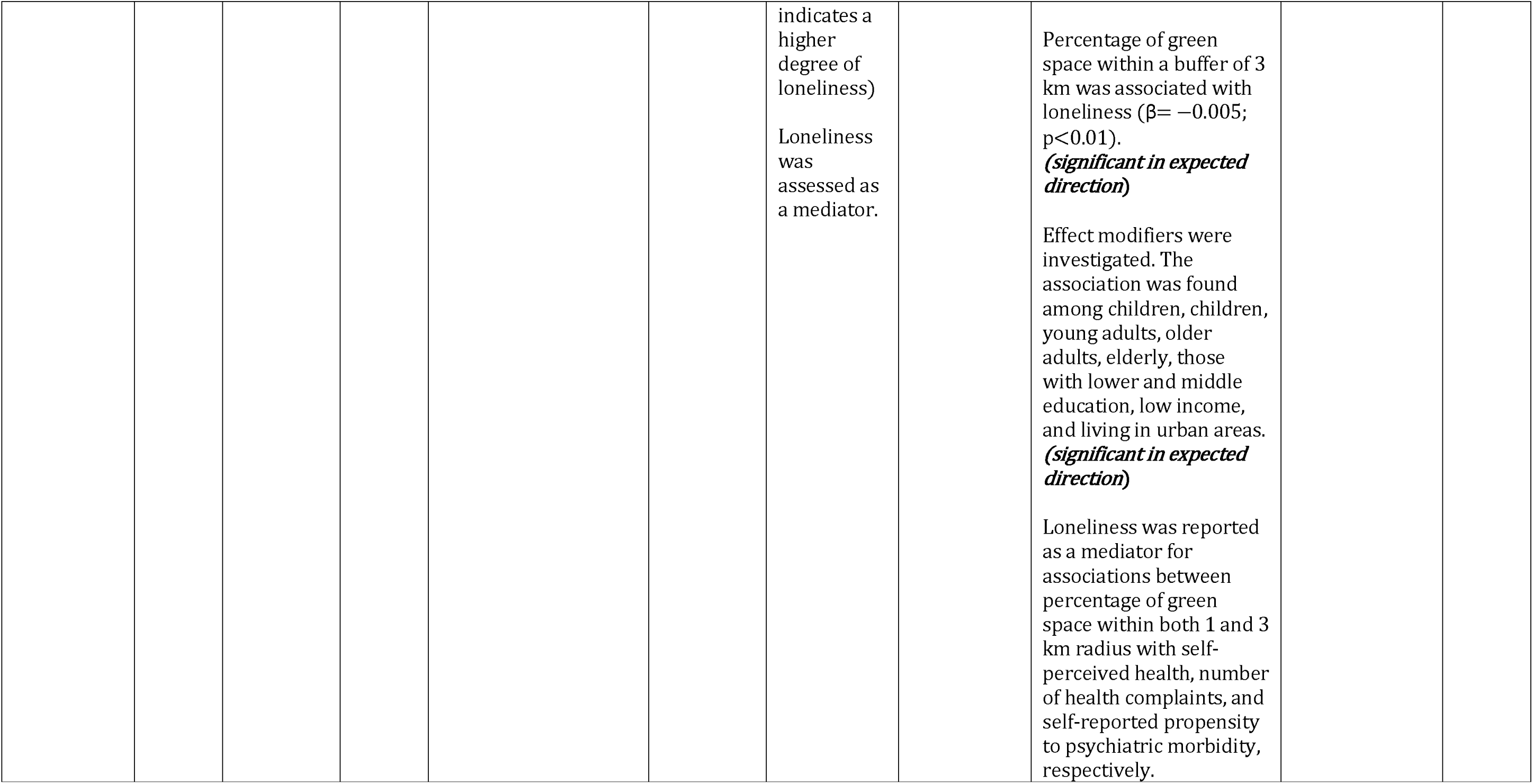
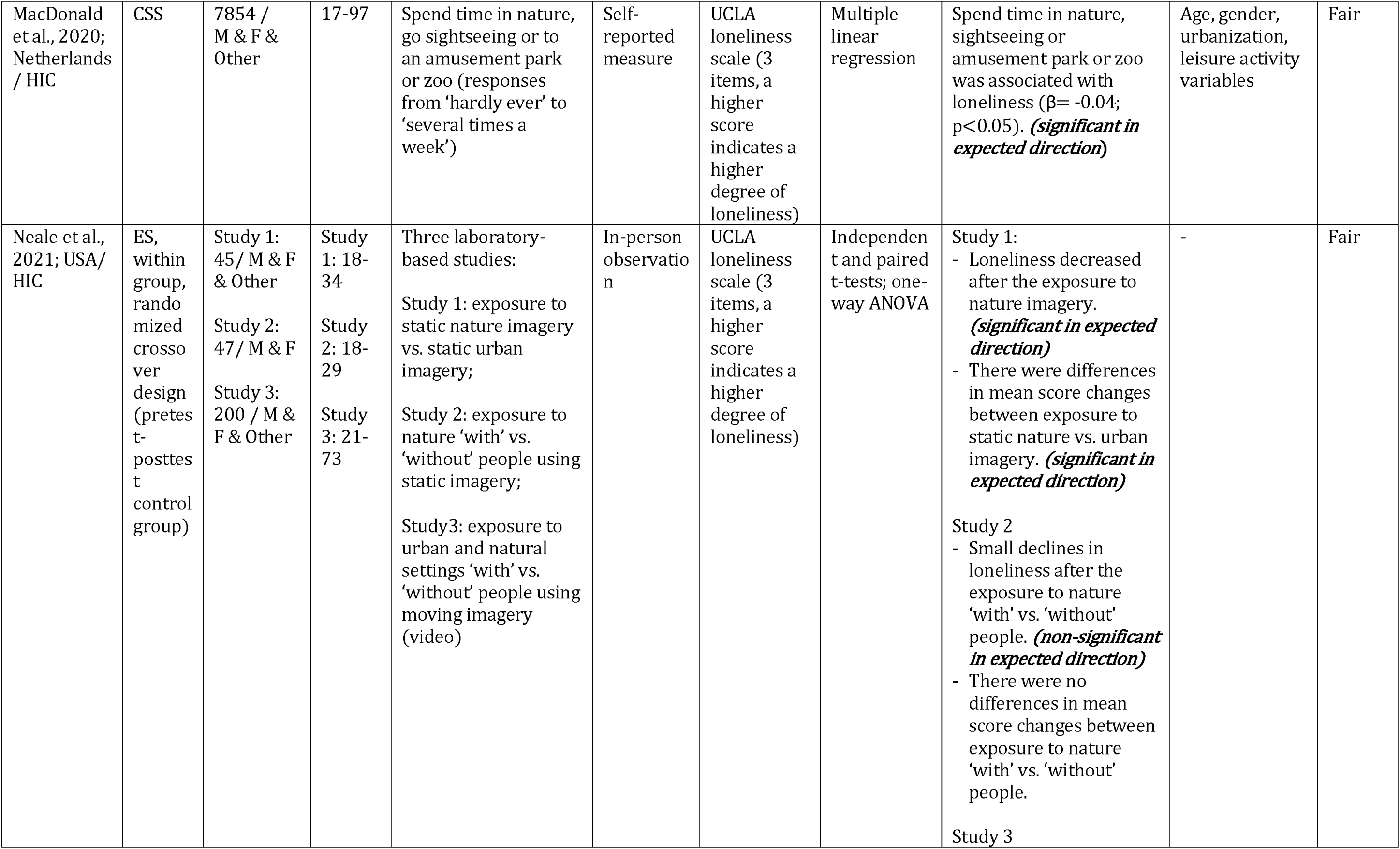
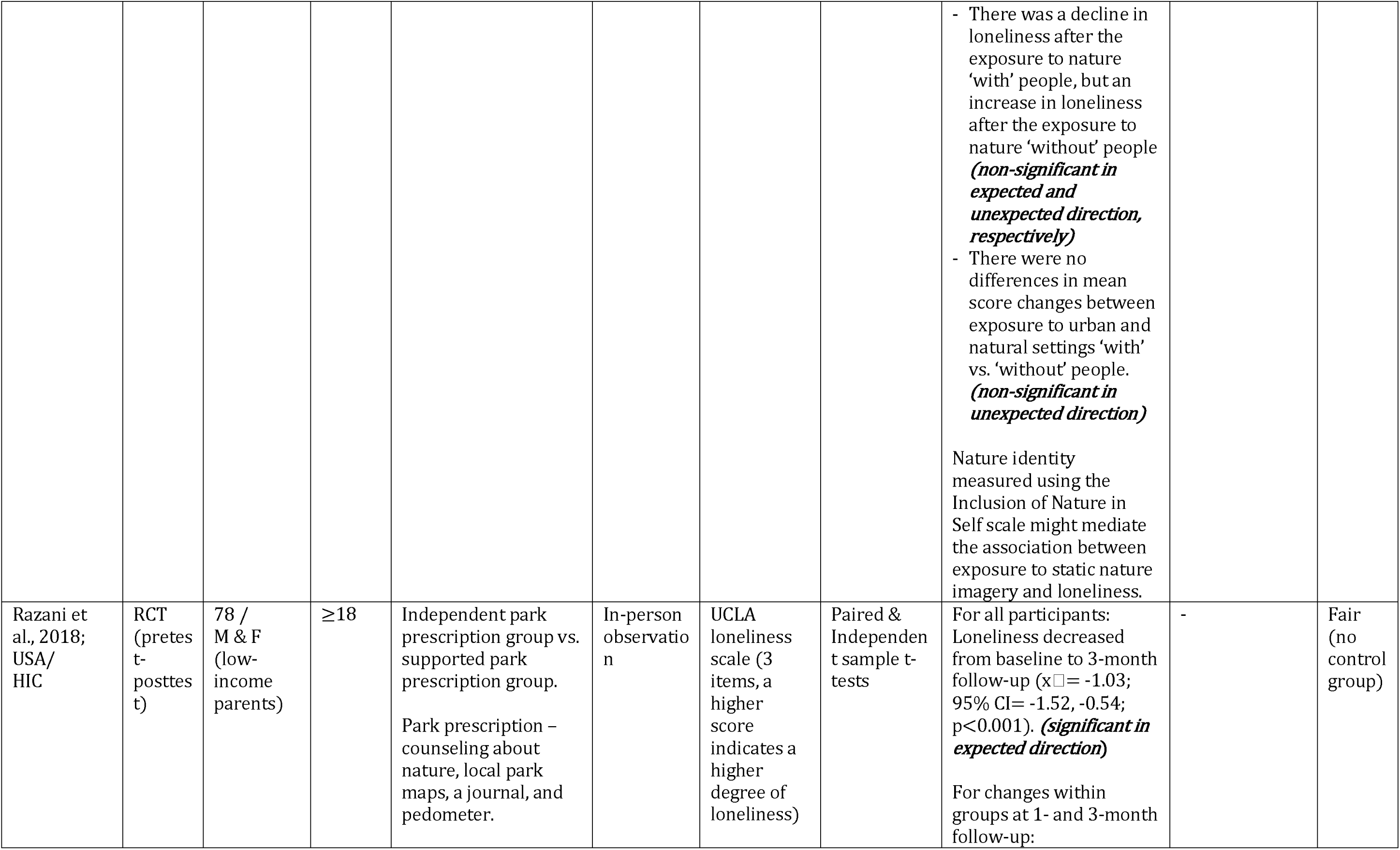
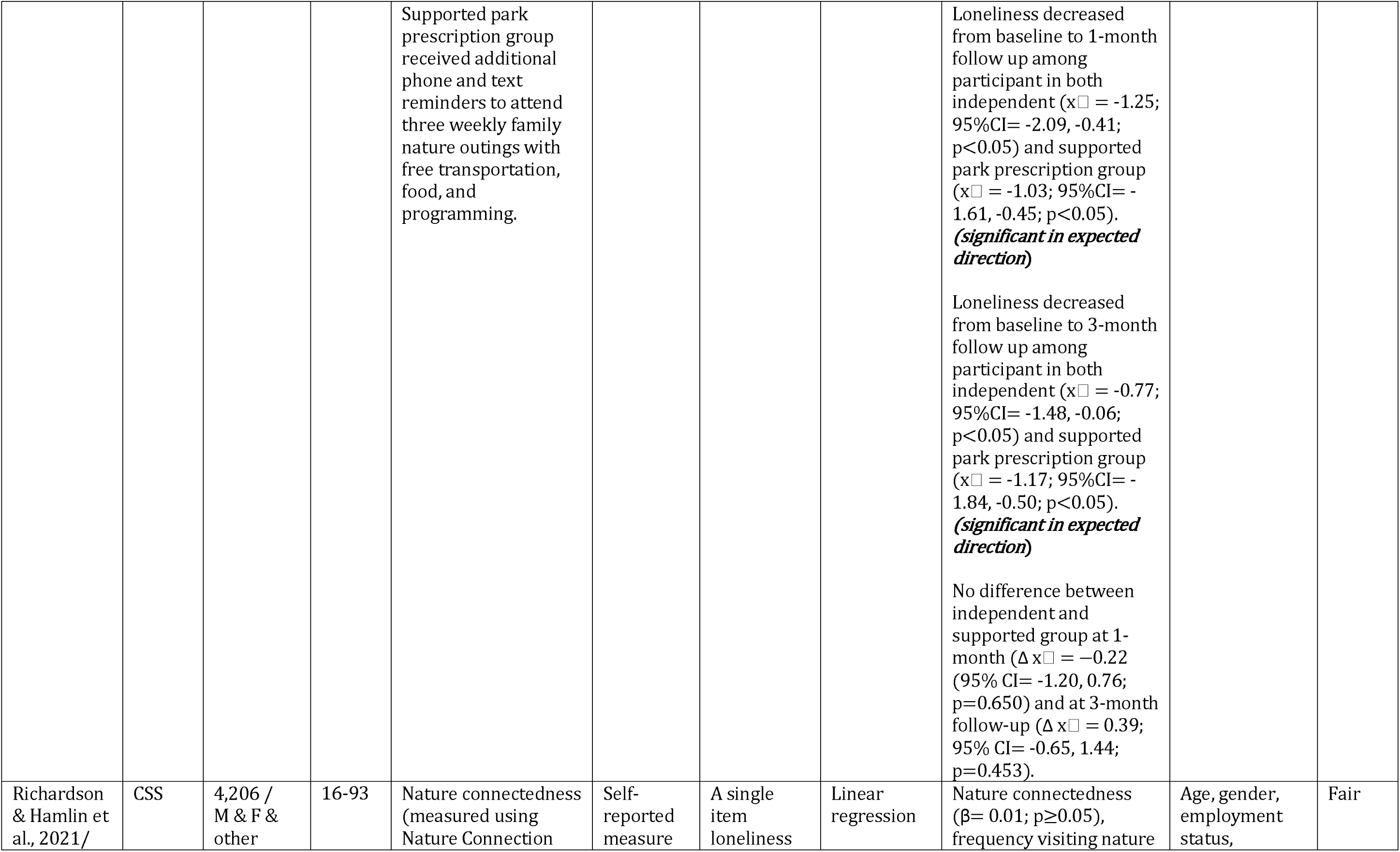
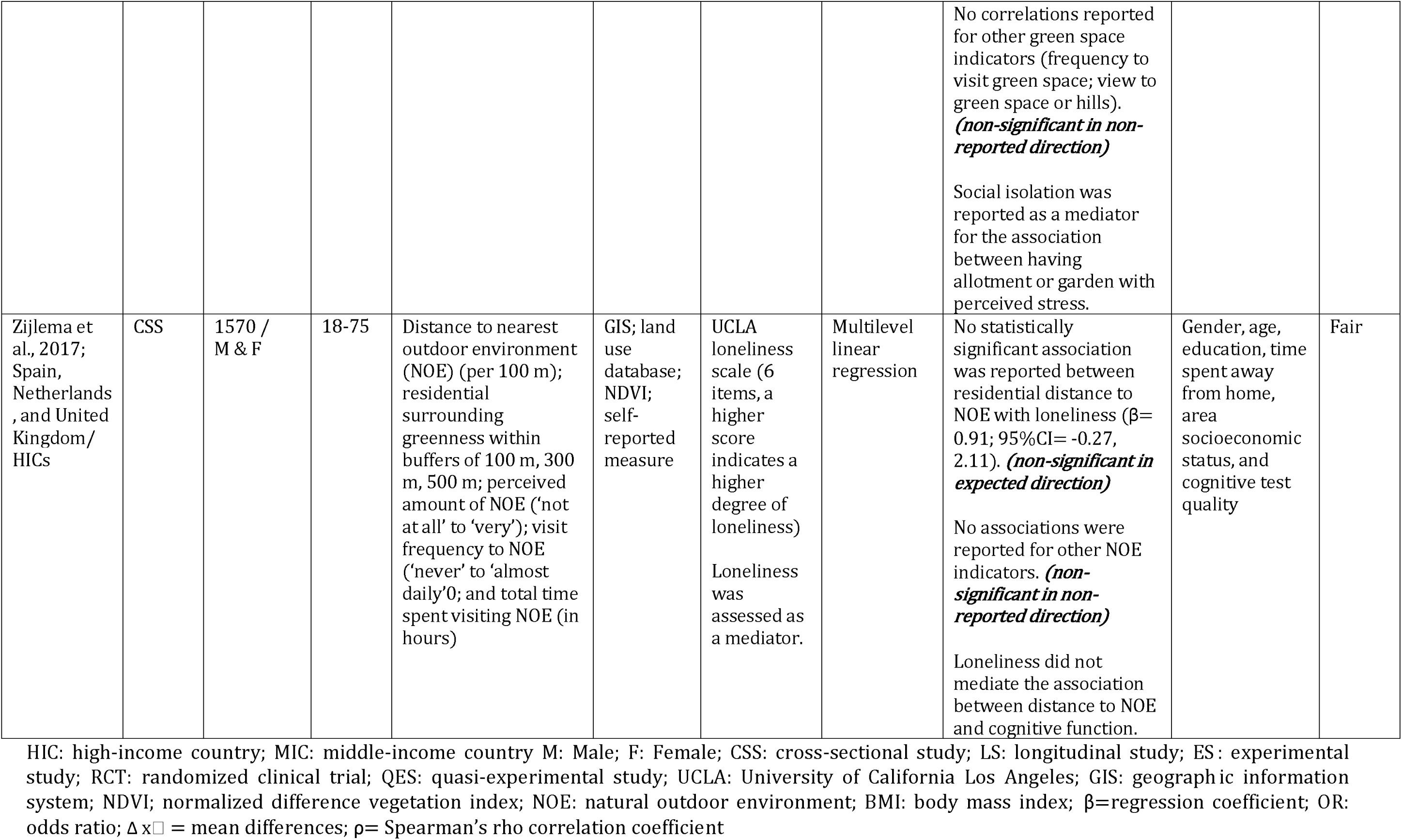
(ONLINE ONLY). Study characteristics and results

